# Is there a sex difference in mortality rates in Paediatric Intensive Care Units: A Systematic Review

**DOI:** 10.1101/2022.05.27.22275455

**Authors:** Ofran Almossawi, Amanda Friend, Luigi Palla, Richard G. Feltbower, Sofia Sardo-Infiri, Scott O’Brien, Katie Harron, Simon Nadel, Bianca De Stavola

## Abstract

**Introduction:** Mortality rates in infancy and childhood are lower in females than males. However, for children admitted to Paediatric Intensive Care Units (PICU), mortality has been reported to be lower in males, although males have higher admission rates. This female mortality excess for the subgroup of children admitted in intensive care is not well understood. To address this, we carried out a systematic literature review to summarise the available evidence.

Our review studies the differences in mortality between males and females aged 0 to <18 years, while in a PICU, to examine whether there was a clear difference (in either direction) in PICU mortality between the two sexes, and, if present, to describe the magnitude and direction of this difference.

**Methods and analysis:** Any studies that directly or indirectly reported the rates of mortality in children admitted to intensive care by sex were eligible for inclusion. The search strings were based on terms related to the population (those admitted into a paediatric intensive care unit), the exposure (sex), and the outcome (mortality). We used the search databases MEDLINE, Embase, and Web of Science as these cover relevant clinical publications. We assessed the reliability of included studies using a modified version of the risk of bias in observational studies of exposures (ROBINS-E) tool. We considered estimating a pooled effect if there were at least three studies with similar populations, periods of follow-up while in PICU, and adjustment variables.

**Results:** We identified 124 studies of which 114 reported counts of deaths by males and females which gave a population of 278,274 children for analysis, involving 121,800 (44%) females and 156,474 males (56%). The number of deaths and mortality rate for females were 5,614 (4.61%), and for males 6,828 (4.36%). In the pooled analysis, the odds ratio of female to male mortality was 1.06 [1.01 to 1.11] for the fixed effect model, and 1.10 [1.00 to 1.21] for the random effects model.

**Conclusion:** Overall, males have a higher admission rate to PCU, and a lower overall mortality in PICU.

**Systematic review registration:** PROSPERO database reference number CRD42020203009.

## 1 Introduction

Child mortality is a global measure of a nation’s health and a top priority for the UK health system^1^. Differences in child mortality rates between the sexes are well documented in almost all developed countries, showing higher female survival rates than males^2^. Overall childhood mortality is very low in the UK, and in other developed countries (United Nations Inter-agency Group for Child Mortality Estimation (2021)). Office for National Statistics (ONS) figures show downward mortality trends in the UK for both males and females since the 1950’s, and levelling off since 2010.

Paediatric Intensive Care Unit (PICU) deaths account for about 15% of all UK childhood fatalities^3^ and 86% of UK hospital deaths^4^ thus provide a sizeable population to study childhood deaths. This led to the design and implementation of a longitudinal study of all infants admitted to UK PICUs over 11 years, which showed a higher PICU mortality rate for female over male infants^5^. This difference is in the opposite direction to that seen in the overall population and could be due to differences in severity of disease on admission, despite both sexes having the same mean and median Paediatric Index of Mortality (PIM2), a proxy for severity of disease at the time of admission and mortality risk score. There are a number of published studies showing similar conclusions but there is no published systematic review which has collated and evaluated all the available evidence.

The aim of this systematic review was to study the differences in mortality, in either direction, between males and females from age 0 to <18 years, where the death event happens in PICU. This review is also part of a wider project using linked PICU and Hospital Episode Statistics (HES) data which aims to study differences in sex mortality and long term outcomes in England^6^.

### 1.1 Aims and Objectives

Using published data, our primary aim is to estimate the difference in mortality rates between males and females who die in PICU. This is to identify if male or female sex is associated with differences in mortality rates in PICU.

Our secondary aim is to quantify the rates of admission to PICU for males and females.

Our specific objectives are to report on the evidence with regards to:

- The difference (absolute or relative, as available) in sex mortality in PICU for all children aged 0 to any age <18 years, overall and separately by age groups
- The rates of admission to PICU for all children aged 0 to any age <18 years by sex
- The evidence summarised overall and by any primary diagnostic groups (sub-populations of PICU)

### 1.2 Review Question

- **Population** Children of any age range <18 years old, and admitted to a Paediatric Intensive Care Unit
- **Exposure** Sex
- **Comparison** Comparing male and female mortality rates and their rates of admission to PICU
- **Outcome** Death within a Paediatric Intensive Care Unit

## 2 Methods

Our protocol was reported previously^7^ using the Preferred Reporting Items for Systematic Reviews and Meta-Analyses Protocols (PRISMA-P) guidelines^8^ and registered with the International prospective register of systematic reviews (PROSPERO) database, reference number CRD42020203009.

### 2.1 Information sources and search Strategy

We conducted a systematic search of PubMed, Embase, and Web of Science using a controlled vocabulary (MeSH) and keywords, without date or language limitations. Our last search update was on 20th of December 2020 and our peer reviewed search strategy was described in the protocol and is reported in Appendix 1 (Search Terms and Search Results).

We identified any studies that addressed the association between sex and PICU mortality in children, where sex was the primary exposure. Additionally, we identified all studies where PICU mortality was reported by sex, or where sex was used as a variable for statistical adjustment in the estimation of mortality rates in PICU. We did report but did not pool any estimate reported if sex was a variable for adjustment. This was to ensure we avoided the ‘Table 2 fallacy’, where effect estimates for any of the adjustment variables included in a regression model alongside the main exposure variable cannot be interpreted^9^.

The search strings were based on terms related to the population (children in intensive care), the exposure (sex), and the outcome (in-PICU mortality).

### 2.2 Study Outcomes

The primary outcome is mortality in PICU by sex. Secondary outcomes are rates of admission to PICU, and length of stay in PICU, by sex.

### 2.3 Eligibility and inclusion criteria

Eligibility and inclusions criteria are presented in Table 1.

**Table 1:** The study eligibility criteria following the Population Exposure Comparison and Outcome model

| PECO | Inclusion Criteria | Exclusion Criteria |
| --- | --- | --- |
| Population | Children 0 to any age <18 years admitted to PICU | Studies with premature neonates or focusing on Very Low Birth Weight infants<br>Studies exclusive to neonatal intensive care<br>Studies with mixed adult and paediatric populations where the paediatric results are not separable from the adult results |
| Exposure | Sex used as a primary exposure for mortality<br>Sex reported as a summary statistic or used as covariate for adjustment | Sex not used as a grouping variable for mortality<br>Sex as primary exposure or covariate for adjustment in the analysis of non-mortality outcomes |
| Comparison | Comparing male to female mortality | Comparing categories of variables other than sex |
| Outcomes | Primary: Mortality in PICU | Mortality in PICU not reported |

We included any observational study, clinical trial, or re-analysis of a clinical trial.

### 2.4 Study exclusion criteria

After the eligibility screening, we further scrutinised studies for any of the exclusion criteria listed in Table 1, and some additional criteria listed below.

Studies meeting at least one of the exclusion criteria were excluded as detailed in the full PRISMA flow diagram in Figures 2a and 2b. Specifically, we excluded:

**Figure 1.**
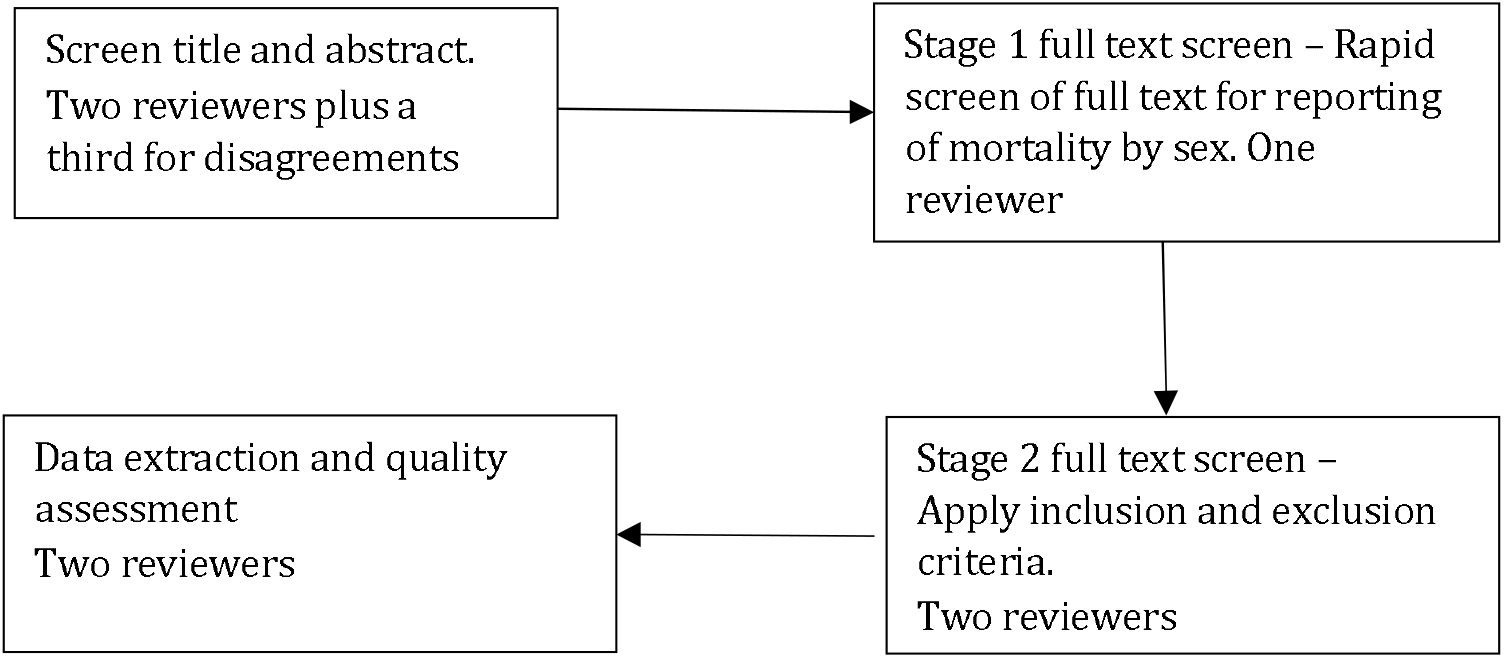
Study screening flow

**Figure 2a:**
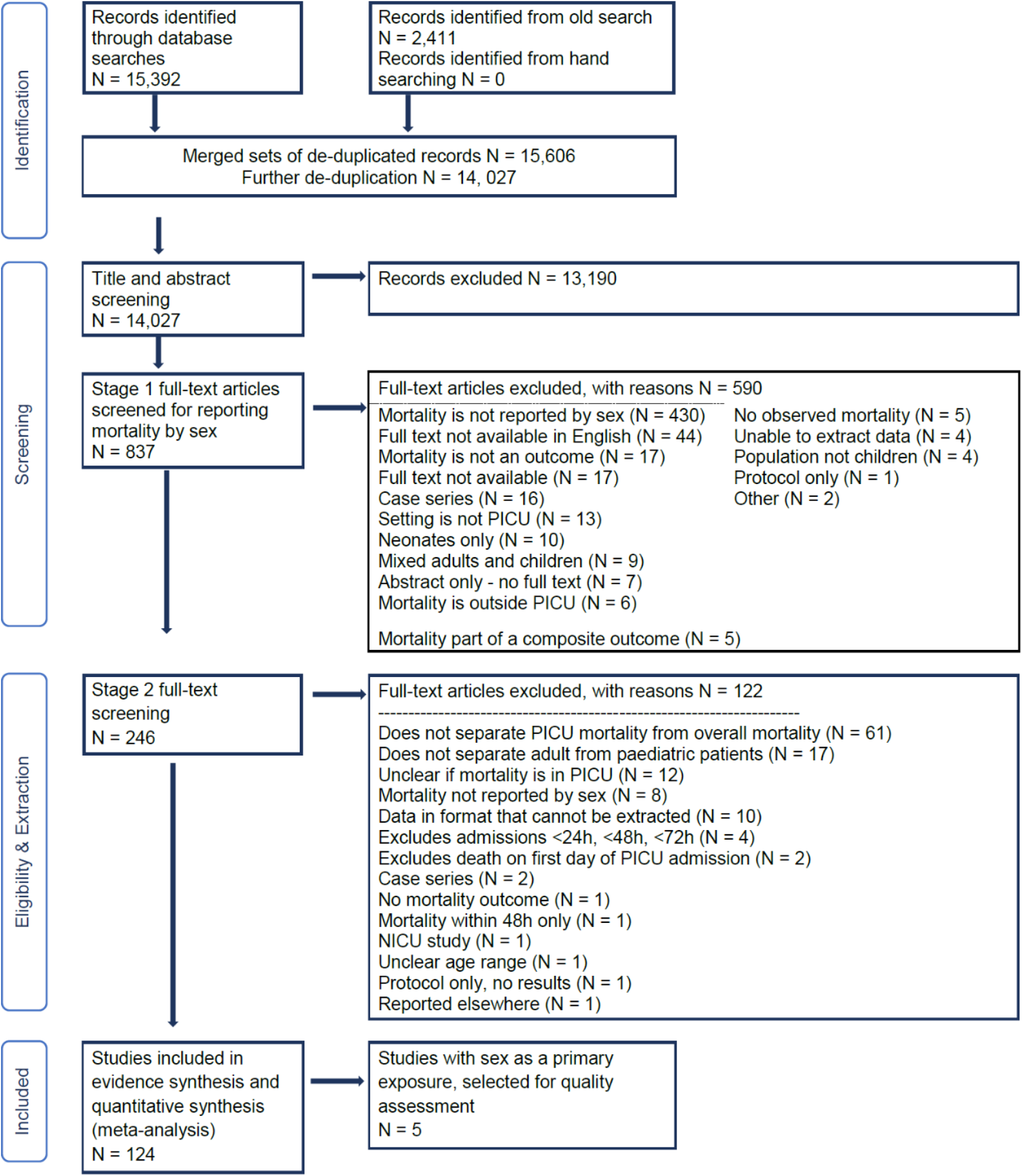
PRISMA flowchart Records identified from the old search are detailed in Appendix 1

**Figure 2b.**
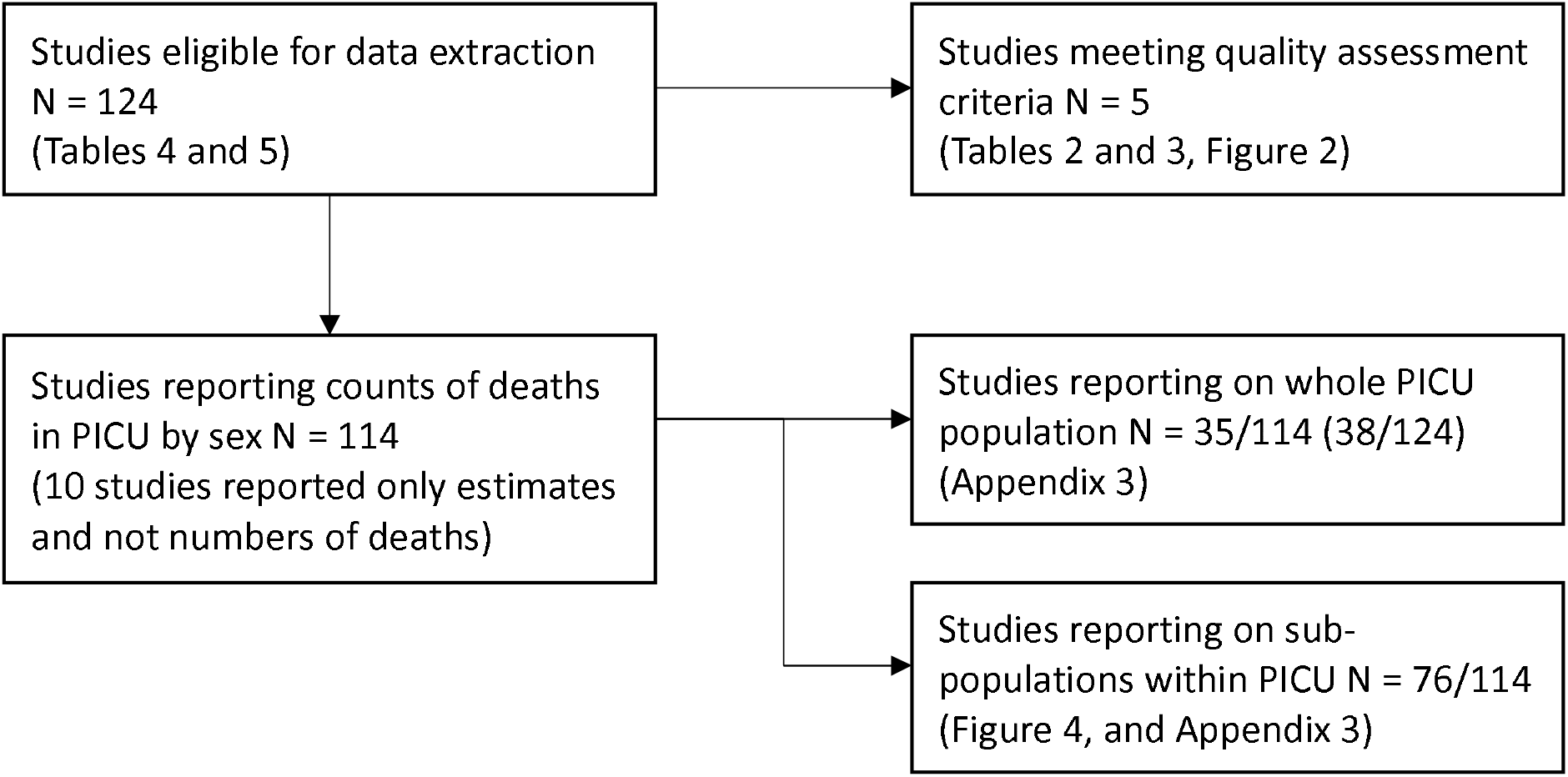
Supplement to PRISMA flowchart

- Studies that were only published in abstract form, or were review articles.
- Potentially, studies not available in English, depending on the a priori specification to exclude non-English language studies if they comprised less than 20% of the full text records.

### 2.5 Study screening mode

#### Screening studies: title and abstract screening

One reviewer screened the titles and abstracts of records after deduplication, and a second reviewer independently checked all the studies from this stage that were labelled ‘yes’ and ‘maybe’ and a sample of the ones labelled as ‘no’. The ‘no’ sample was assigned to be twice the number of the ‘yes’ total. A third reviewer resolved any disagreements. If all three reviewers gave different answers (Yes/No/Maybe) then the study was included.

#### Screening studies: applying inclusion and exclusion criteria

For the studies included at the title and abstract level, we applied full text screening in two stages. Stage 1 was a rapid screening carried out by one reviewer to verify if the mortality outcome was reported by each sex. Stage 2 was applied to the studies included from stage 1, where we applied the remaining inclusion and exclusion criteria and this was done by two reviewers independently. See Figure 1.

#### Screening studies: quality assurance process

The inclusion/exclusion decisions made by the reviewers on the basis of titles and abstract were compared and agreement summarised using kappa statistics. We calculated the level of agreement between rates at this stage using Cohen’s weighted kappa. We used weights that reflected a disagreement of ‘maybe/yes’ or ‘maybe/no’ carries less weight than ‘yes/no’.

### 2.6 Critical appraisal and data extraction

We adapted the DistillerSR software^10^ for data extraction to capture specific features for our study. The resulting tool was piloted and rectified before full extraction was performed by one reviewer. Two additional reviewers independently checked the extracted data. The full data extraction sheet and risk of bias tool are available in Appendix 2 (Tools used in screening, extraction, and quality assessment).

Studies where sex was the main exposure of interest were eligible for quality assessment using the “risk of bias in observational studies of exposures” (ROBINS-E) tool^11,12^, which scores studies to be of high, unclear and low risk of bias. Two reviewers independently assessed and checked eligible studies for quality, while a third reviewer resolved any disagreements between the first two reviewers.

### 2.7 Data analysis and synthesis

We carried out a narrative synthesis of the data, with two final summary tables. The first is for studies with sex as the main exposure of interest, and the second is for all studies, including those where sex was used as a variable for adjustment or a variable for summary statistics.

Where we had three or more studies with a similar sub-population e.g. admissions due to sepsis, we present their results graphically in a forest plot. As a summary report, we combined all studies with death numbers reported by sex, regardless of their variability and types of sub-populations.

We categorised the reported age groups to enable pooling of some studies that have a similar population and with the same age group, see Table 2.

**Table 2.**
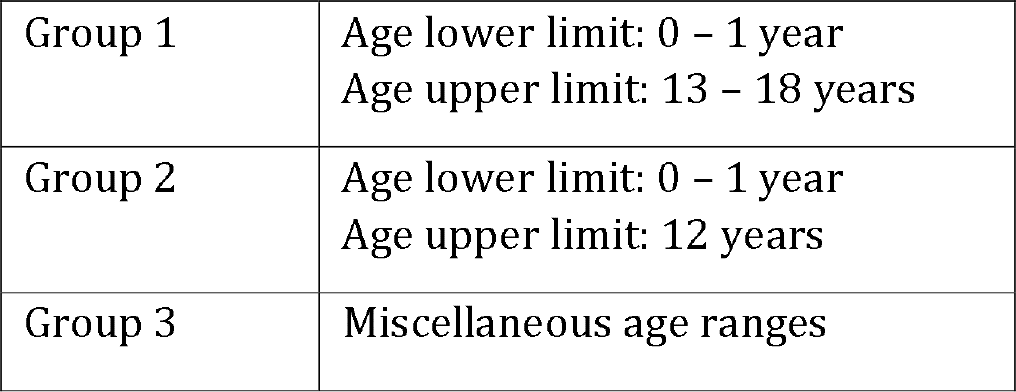
Age groups for the included studies

|  |  |
| --- | --- |
| Group 1 | Age lower limit: 0 – 1 year<br>Age upper limit: 13 – 18 years |
| Group 2 | Age lower limit: 0 – 1 year<br>Age upper limit: 12 years |
| Group 3 | Miscellaneous age ranges |

All analyses were carried out in R version 4.1.1.

### 2.8 Protocol changes

In our protocol we planned to summarise mortality after PICU discharge in addition to mortality in PICU. However, after summarising the variability in the studies, we concluded that additional information on out of PICU mortality would not confer additional knowledge due to the variability in the reporting of post-PICU mortality.

## 3 Results

Our search strategy identified 15,392 studies, of which 124 were eligible for inclusion, see Figure 2a. Overall, the 124 included studies had a total population of 866,620children, 379,733 (44%) females and 486,887 (56%) males. Of the 124 studies, 114 reported counts of deaths by males and females which give a population of 278,274 children for analysis, specifically involving 121,800 (44%) females and 156,474 males (56%). The number of deaths and mortality rate for females was 5,614 (4.61%), and for males 6,828 (4.36%); thus there is a slightly higher proportion of deaths in females.

One reviewer screened the titles and abstracts of 14,028 studies, and a second reviewer blindly double checked all the included studies (Yes = 863, Maybe = 406) from this stage and a sample of the excluded ones, totalling 2,562 double checks. The level of agreement and weighted Kappa was 68.7% and 0.62 respectively. This was driven mostly by the answers being yes/no/maybe, where a ‘maybe’ answer was given if the abstract mentioned sex as a variable, but did not make clear if the mortality outcome was reported for each sex. This was also reflected in our exclusion reasons in Figure 2a, where we excluded 430 records out of 837 due lack of mortality numbers by sex. When we excluded the ‘maybe’ records, the level of agreement and kappa were 88.5% and 0.69.

We were unable to retrieve the full text of 17 articles, and did not scrutinise the full text of the non-English articles. The non-English records were 44 out of 837 (5.3%) therefore excluded as they comprised <20% of the full text records eligible for screening. We retrieved the full text for the remaining 776 studies and applied the exclusion criteria in two stages. In stage 1, one reviewer rapidly assessed if the mortality outcome was reported by sex. In stage 2, a reviewer applied the exclusion criteria to the remaining 246 studies, and a second reviewer checked this process. The remaining 124 studies were eligible for data extraction. See Figures 2a and 2b for full details.

### 3.1 Tables of study summaries

We report two types of summaries: first for all the studies meeting our extraction criteria (N = 124), and then for the subset of these studies where sex was the main exposure of interest and for which mortality was reported separately by sex (N = 5), see Table 3. To simplify the reporting, we split the summary of the 124 studies into two parts depending on the mortality outcomes for males and females, see Appendix 3 (Summary tables of 124 studies meeting the inclusion criteria)

**Table 3.**
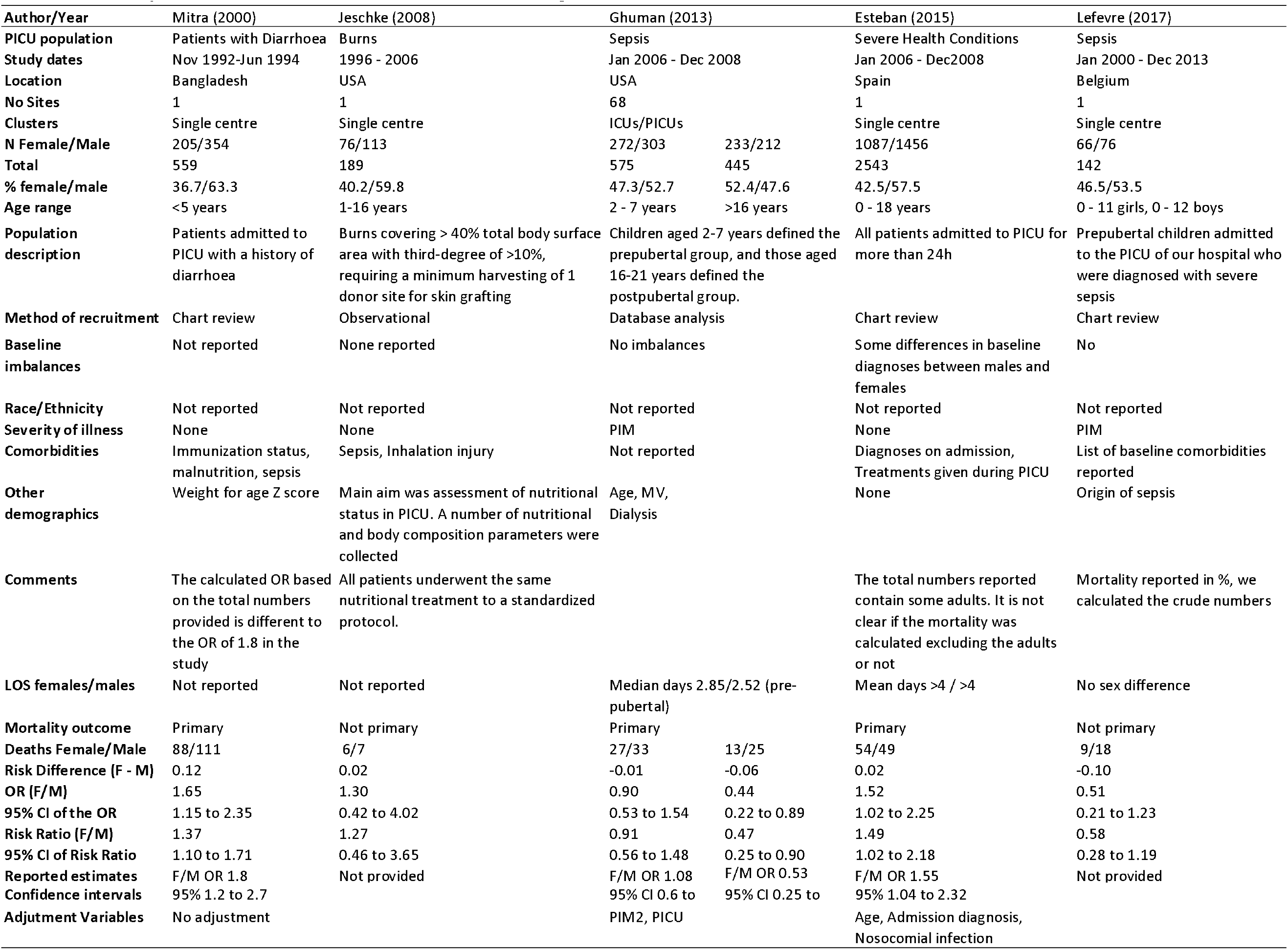
Summary of the five studies where sex was the main exposure

We report the measures of association between sex and mortality in two ways. If the crude numbers of deaths were reported by sex, we calculated the measure of association in terms of odds ratios. Otherwise, we present the reported measure of association and list any adjustment variables if used.

We report all the measures of association along with their confidence intervals (CIs), the type of sub-population, the age group, and the set of adjustment variables if used in each study. Only 18 of the 124 studies reported a measure of association of sex on mortality. All other studies reported numbers of deaths by sex as a summary statistic, see Appendix 3 (Summary tables of 124 studies meeting the inclusion criteria). To summarise the results presented in these two tables, 68 studies reported higher female mortality, 6 studies reported equal mortality, and 50 studies reported higher male mortality.

### 3.2 Sex as the main exposure

Overall we found eight studies addressing sex as the primary exposure. Of these eight, three were excluded because PICU mortality was not reported separately from other mortality outcomes^13–15^.

Table 3 summarises the five studies that met our criteria for quality assessment. There is considerable variability between these studies in terms of the age range, sub-population of PICU and baseline characteristics such as co-morbidities. Four of these studies did not include any score for severity of disease on admission; one reported the Paediatric Index of Mortality (PIM) score. Although all five studies specified sex as the primary exposure, in two of them PICU mortality was not the primary outcome. All studies reported a lower percentage of female admissions compared to males.

When we used the crude numbers to calculate the association between sex and mortality, three of the studies showed higher female mortality relative to males. In one of the two papers where male mortality was higher, the adjusted association reported by the authors showed the opposite, see Ghuman^16^.

Table 4 shows the quality assessment of the five studies using a modified version of the ROBINS-E tool. None of the studies achieved a high score for quality.

**Table 4.** Quality assessment of the five studies where sex was the main exposure, using the ROBINS-E tool

| Author | Mitra<sup>17</sup> | Jeschke<sup>18</sup> | Ghuman<sup>16</sup> | Esteban<sup>19</sup> | Lefevre<sup>20</sup> |
| --- | --- | --- | --- | --- | --- |
| Year | 2000 | 2008 | 2013 | 2015 | 2017 |
| Country | Bangladesh | USA | USA | Spain | Belgium |
| Exposed/Non Exposed Same Population | Probably yes | Definitely yes (low risk of bias) | Definitely yes (low risk of bias) | Probably yes | Definitely yes (low risk of bias) |
| Confidence Of Assessment Of Exposure | Definitely yes (low risk of bias) | Definitely yes (low risk of bias) | Definitely yes (low risk of bias) | Definitely yes (low risk of bias) | Definitely yes (low risk of bias) |
| Confident Outcome Not Present At Start | Definitely yes (low risk of bias) | Definitely yes (low risk of bias) | Definitely yes (low risk of bias) | Definitely yes (low risk of bias) | Definitely yes (low risk of bias) |
| Adjusted For Baseline Variables | Definitely no (high risk of bias) | Mostly yes | Mostly yes | Mostly yes | Mostly yes |
| Assessment Presence/Absence Baseline Variables | Probably no | Probably yes | Probably yes | Probably yes | Probably yes |
| Assessment Of Outcome | Definitely yes (low risk of bias) | Definitely yes (low risk of bias) | Definitely yes (low risk of bias) | Definitely yes (low risk of bias) | Definitely yes (low risk of bias) |
| Follow up Cohorts Adequate | Probably yes | Definitely yes (low risk of bias) | Definitely yes (low risk of bias) | Probably yes | Probably yes |
| Group Interventions Similar | Probably yes | Probably yes | Probably yes | Probably yes | Probably yes |
| **Assessment of Bias** | High risk of bias for one or more key domains. | Unclear risk of bias for one or more key domains. | Unclear risk of bias for one or more key domains. | Unclear risk of bias for one or more key domains. | Unclear risk of bias for one or more key domains. |

### 3.3 Sex as a baseline variable

In addition to the five studies where sex was the primary exposure, we summarised the results for a further 119 studies where the numbers of deaths for each sex were reported as a summary statistic, or sex was used as a variable for adjustment when studying mortality in PICU and estimated associations were reported for it. Appendix 3 (Summary tables of 124 studies meeting the inclusion criteria)

### 3.4 Other secondary outcomes

Proportions of PICU admission by sex are reported in Appendix 3 (Summary tables of 124 studies meeting the inclusion criteria). Out of 124 studies, 14 (11%) reported higher proportion of female admissions. However, the study by Ghuman^16^ reported on two age ranges showing a slightly higher admission rate for females compared to males in the 16 to 21 years age category relative to younger ages. As the former group is a mixture of adults and paediatric patients, it fell outside the criteria of inclusion for this review.

For the length of stay outcome, 118 studies did not report this outcome by sex. For the five studies meeting the quality assessment, we have reported a summary of this outcome in Table 2.

### 3.5 Variability in sub-populations

We found wide variability between the studies with regards to the sub-populations of PICU and their age range. It was therefore difficult to combine the results. Figures 3 and 4 summarise the numbers and proportions of population types we found in the studies which are summarised in Table 3 and Appendix 3 respectively.

**Figure 3.**
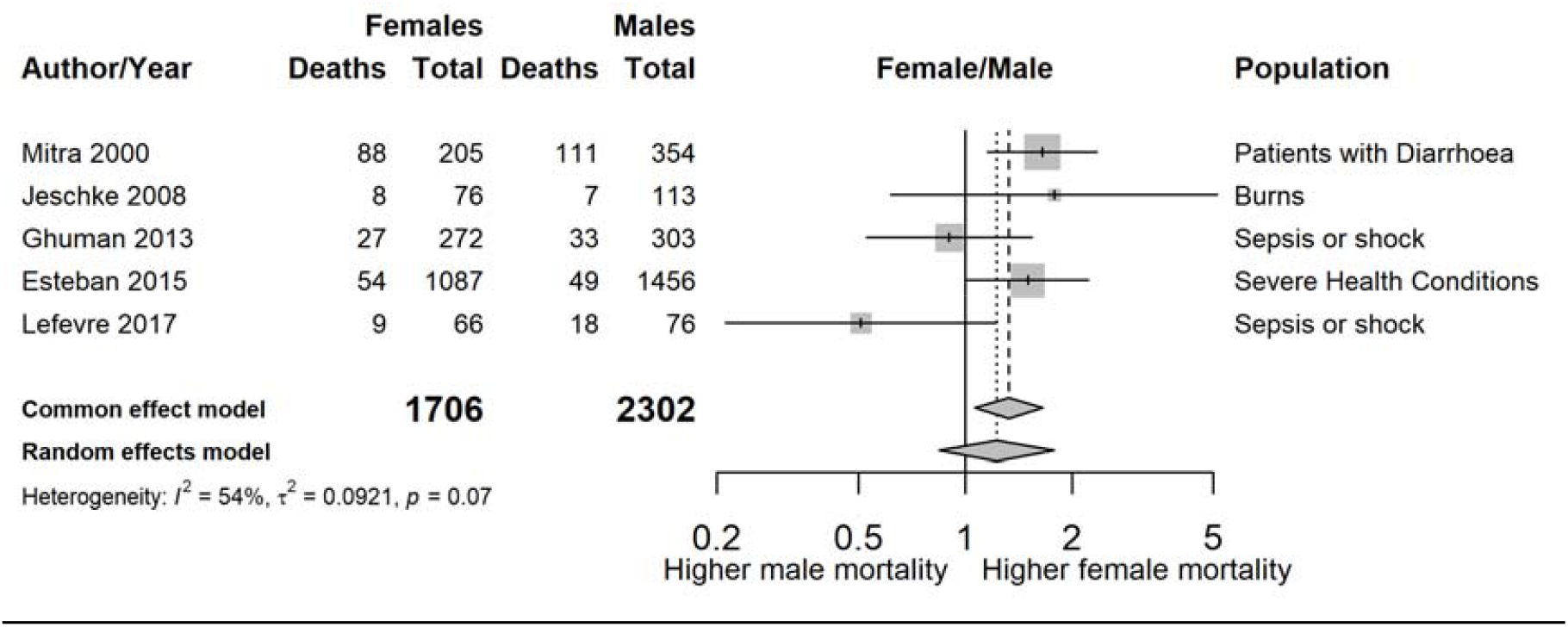
Forest plot showing the estimated unadjusted odds ratios of female to male mortality by study, sorted by year of publication

**Figure 4.**
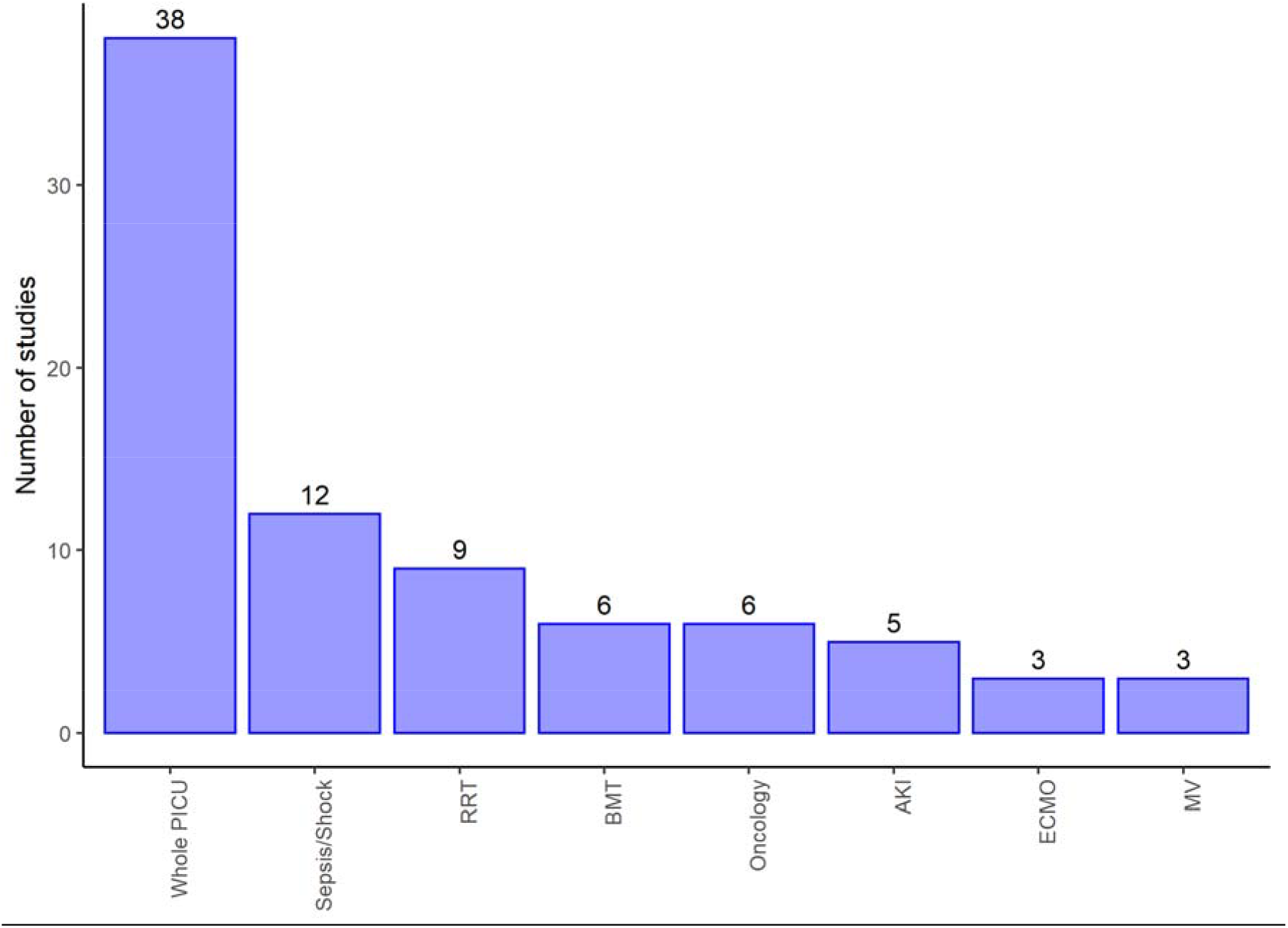
Number of studies by type of PICU admission of the reported studies summarised in Appendix 3 Displays populations reported by at least three of the studies selected for extraction and make up 82/124 (66%) of these studies, and 72/124 (58%) reported counts of death by sex RRT: Renal replacement therapy; BMT: Bone marrow transplant; AKI: Acute kidney injury; ECMO: Extra corporeal membrane oxygenation; MV: Mechanical ventilation

### 3.6 Publication bias

As far as we could assess, we found very little evidence for publication bias in the reporting of studies. Figure 5 shows a funnel plot of the 28 studies of whole PICU population categorised into age group 1, showing negligible asymmetry. We focus on this subgroup of results because they should be more homogeneous in effect estimates.

**Figure 5.**
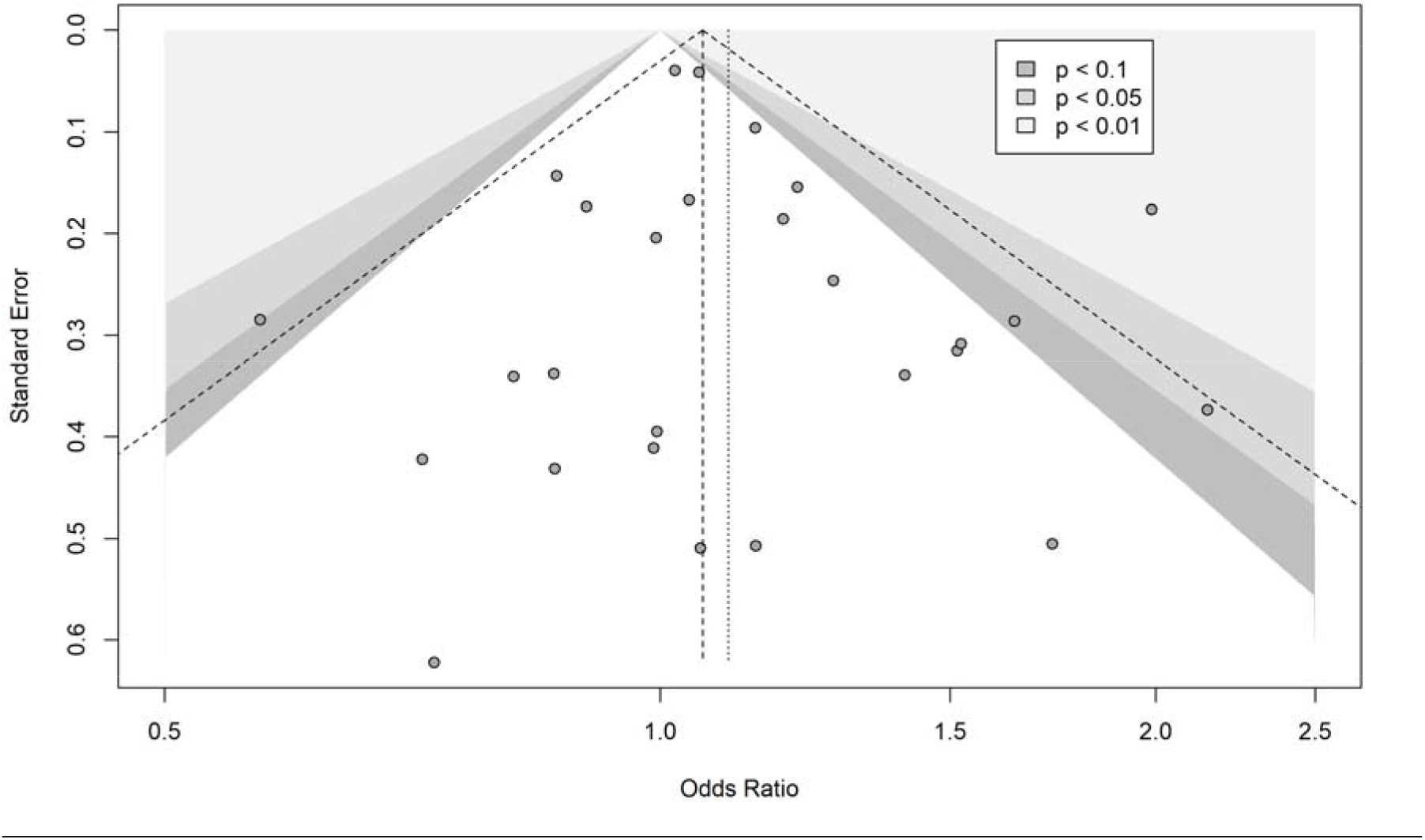
Funnel plot of 27 studies reporting on whole PICU population and belong to age group 1

### 3.7 Summary of studies reporting counts of death

Figure 3 shows a summary plot of the crude odds ratios for the five studies where sex was the primary exposure. We have not combined the estimates due to the large variability (I^2^ = 53.6% [0.0% to 82.9%]) in sub-populations and age ranges between the studies.

From the remaining 119 studies that do not meet the quality assessment criteria, we report a summary plot of the estimated odds ratios of female to male mortality for the 27 studies which included whole PICU populations in age group 1 (see Figure 6). The unadjusted pooled OR of female to male mortality is 1.06 for the common (i.e. fixed) effect model, and 1.10 for the random effects model, with no strong evidence of heterogeneity (I^2^ = 29%).

**Figure 6.**
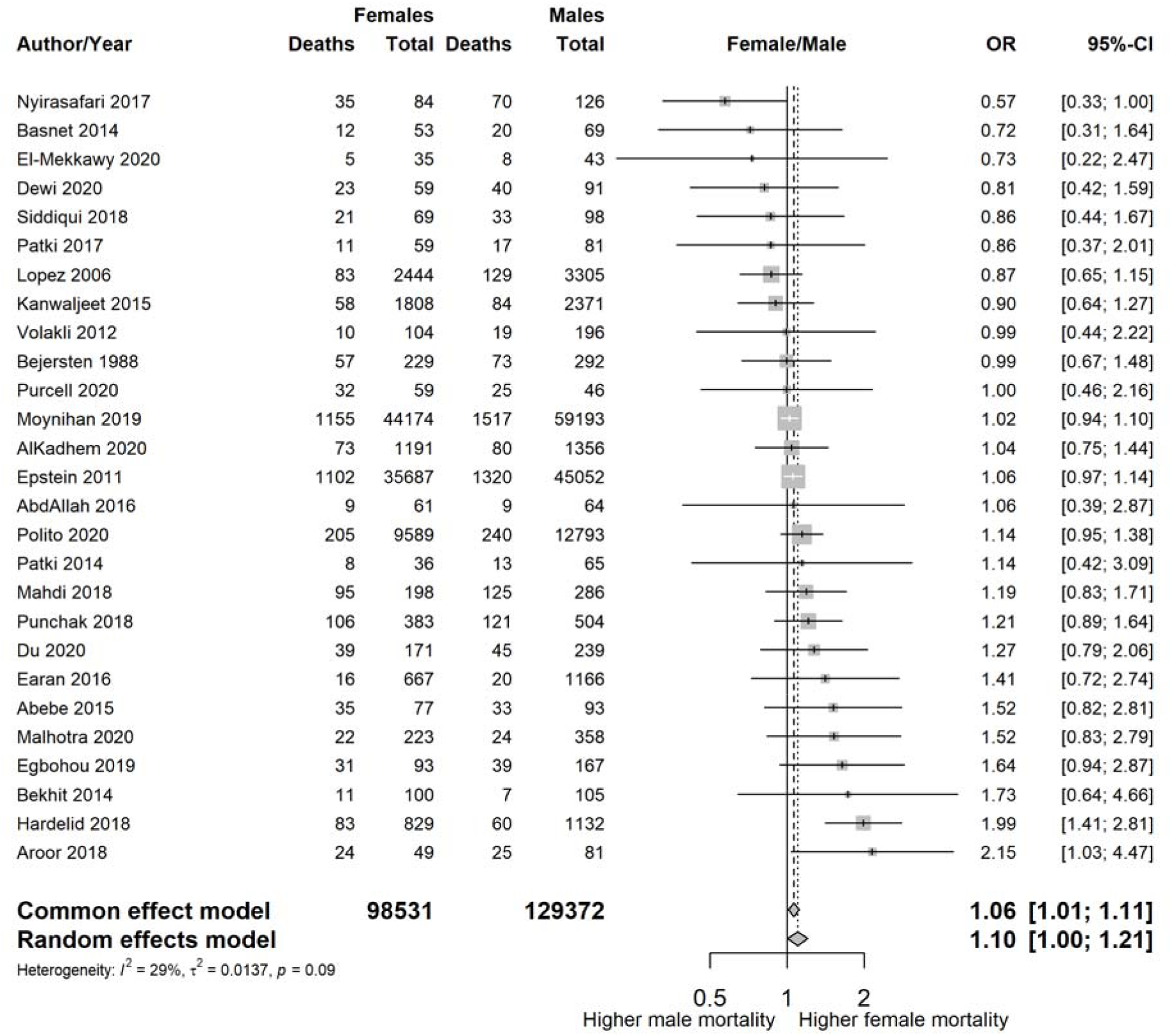
Estimated odds ratios of female to male mortality for 27 studies that include the whole PICU population belonging to age group 1, sorted by the magnitude of the odds ratio

Additional plots of sub-populations reported in three studies or more can be found in Appendix 4 (Additional plots for some of the reported sub-populations)

When we combined the 114 studies reporting death counts in a pooled estimate, regardless of their heterogeneity, we had data on 278,274 individuals and 12,442 deaths. The unadjusted pooled OR of female to male mortality was 1.11 [95% CI 1.07 to 1.15] for the common (i.e. fixed) effect model, and 1.14 [95% CI 1.04 to 1.26] for the random effects model. The I^2^ statistic reflecting heterogeneity between studies was 58.9% [95% range 49.9% to 66.6%] with a p value of <0.001, indicating a high degree of heterogeneity. Hence these overall estimates are reported only as an indication of the possible direction of the association.

## 4 Discussion

Our systematic review shows that whilst more male children are admitted to PICU, females tend to be more likely to die in PICU than males. Depending on the study, female mortality rates ranged from lower (OR 0.14) to higher (OR 5.06) than males, with a predominance (55%) of studies reporting higher female mortality. A number of studies (5%) reported similar mortality rates between sexes, in contrast to population mortality rates, where male mortality is higher.

Our review captured a wide range of studies in terms of design, size and variety of PICU sub-populations. This resulted in the full text scrutiny of over 837 studies and the inclusion of 124. However, we were only able to identify eight studies that reported sex as the primary exposure and only five eligible for data extraction. Nevertheless we were able to summarise the findings with a large number of participants, N = 866,620. For the majority of studies (n=119), the publication year was after 2000 reflecting the clinical and reporting progress made in paediatric intensive care data capture over the last two decades.

Another strength of this review is that there appears to be little publication bias since investigating the association between sex and mortality was not the primary aim of the majority of studies.

One of the limitations of our review is that it was not possible to combine the study estimates due to the large variability in the PICU sub-populations analysed, and the age ranges of the children included in these analyses. Where the association between sex and mortality was reported, and adjustments for confounders included, the variables used to statistically adjust the association between sex and mortality widely varied between studies. Studies reporting adjusted estimates for mortality did not justify the selection of variables used for their statistical adjustments and no two studies with adjusted mortality outcomes were comparable.

Furthermore, follow-up periods for reporting death in PICU were variable, with some studies reporting 7-day and 30-day outcomes in addition to the overall mortality. It was not clear if the 30-day outcomes were for deaths occurring in PICU or post discharge from PICU.

Other limitations are that we only considered deaths in PICU, and excluded studies on exclusively neonatal admissions.

We were only able to find five studies, none of good quality, where sex was addressed as the primary exposure. In some of these studies adjustment variables were used, but without rigorous justification for the set of variables used.

These findings show a paucity of evidence in relation to the effect of sex on mortality. Understanding the mechanisms for these differences can assist in improved identification of higher risk children and potentially improvements in the mortality scoring systems used in PICU. A robust and sufficiently large study of PICU mortality in children is needed, where confounder identification and selection is carried out methodically to enable a mechanistic study of the relationship between sex and mortality in PICU.

## 5 Conclusion

The evidence we have collected shows that, among children admitted to PICU, females appear to have a higher risk of PICU mortality than males, in contrast to a male excess of admissions to PICU. Investigating the reasons for these disparities may help improve insights into the needs of specific populations of critically ill children.

The number of children contributing to this review was large but the quality of the reporting studies were average or poor. Pooling of estimates was not possible in general due to their variability in design.

## Supporting information

Appendix 1 (Search Terms and Search Results)

Appendix 2 (Tools used in screening, extraction, and quality assessment)

Appendix 3. Tables of study summaries

Appendix 4 (Additional plots for some of the reported sub-populations

PRISMA checklist

## Data Availability

All relevant data are within the manuscript and its Supporting Information files.

## Supporting information

Appendix 1: Search Terms and Search Results

Appendix 2: Tools used in screening, extraction, and quality assessment

Appendix 3: Summary tables of 124 studies meeting the inclusion criteria

Appendix 4: Additional plots for some of the reported sub-populations PRISMA-P checklist

## 6 Ethics and consent

Ethical approval is not required for this review as it synthesises data from existing studies. This manuscript is a part of a larger data linkage study, for which Ethical approval was granted by the London - City & East Research Ethics Committee, REC reference: 19/LO/1396, IRAS project ID: 214031.

## 8 Availability of data and materials

The datasets used and/or analysed during the current study are available from the corresponding author on reasonable request.

## 9 Patients and public involvement

This review is part of a larger research project with a Project Advisory Group (PAG). Members of the PAG have reviewed this manuscript. Details of the main project can be found on the UCL Child Informatics Group Webpage.

## 10 Authors’ contributions

OA conceived the idea for the literature review and drafted the protocol. BD and RF reviewed and refined the protocol aims and objectives. BD, KH, RF, AF, and LP reviewed, contributed to, and approved the manuscript for the protocol. OA conducted the literature search, AF and LP reviewed the search strategy and approved it. OA, AF, LP and SSI screened all the titles and abstracts and resolved conflicts from the title and abstracts review. For the included full text publications, OA and SOB extracted the data and completed the risk of bias tool then checked these steps. OA conducted the analysis and drafted the manuscript. All authors reviewed and approved the final draft.

## 11 Acknowledgements

We would like to thank members of the Project Advisory Group, Paul Saunders and Viki Ainsworth, for their contributions to the larger project and this manuscript in particular. We are grateful to our funder, the National Institute for Health Research, for their fellowship grant to Ofran Almossawi. This research was supported in part by the NIHR Great Ormond Street Hospital Biomedical Research Centre.

## 12 List of Abbreviations

HES: Hospital Episode Statistics. 2
ONS: Office of National Statistics. 2
PICU: Paediatric Intensive Care Unit. 1–6, 8, 10, 22–25
PIM: Paediatric Index of Mortality. 10
PRISMA-P: Preferred Reporting Items for Systematic Reviews and Meta-Analyses Protocols. 3
R&D: Research and Development. 38

