## Appendix 1 (Search Terms and Search Results) for "Is there a sex difference in mortality rates in Paediatric Intensive Care Units: A Systematic Review"

The search strategy was refined over a number of steps. Table 7A was taken as the main search initially and titles were reviewed. However, after an initial peer review the search was widened as detailed in Table 7C.

**Table 1. Starting search strategy**

| Database | Search terms | | No. of records |
| --- | --- | --- | --- |
| Ovid MEDLINE^®^  1946 to February Week 4 2020 | **1** | mortality/ or child mortality/ or hospital mortality/ or infant mortality/ or survival rate/ | 270402 |
|  | **2** | death.mp. or Death/ or Infant Death/ | 700335 |
|  | **3** | 1 or 2 | 907896 |
|  | **4** | intensive care.mp. | 148776 |
|  | **5** | critical care.mp. or Critical Care/ | 64774 |
|  | **6** | exp pediatric intensive care unit/ | 21366 |
|  | **7** | 4 or 5 or 6 | 183237 |
|  | **8** | child*.mp. or Child/ | 2246889 |
|  | **9** | gender.mp. | 274878 |
|  | **10** | sex.mp. or Sex/ | 754118 |
|  | **11** | 9 or 10 | 934753 |
|  | **12** | 3 and 7 and 8 and 11 | 457 |
| Ovid Embase  1980 to 2020 Week 52 | **1** | mortality/ or child mortality/ or hospital mortality/ or infant mortality/ or survival rate/ | 958289 |
|  | **2** | death.mp. or Death/ or Infant Death/ | 1194930 |
|  | **3** | 1 or 2 | 1943351 |
|  | **4** | intensive care.mp. | 336931 |
|  | **5** | critical care.mp. or Critical Care/ | 124250 |
|  | **6** | exp pediatric intensive care unit/ | 5520 |
|  | **7** | 4 or 5 or 6 | 349453 |
|  | **8** | child*.mp. or Child/ | 2464213 |
|  | **9** | gender.mp. | 558247 |
|  | **10** | sex.mp. or Sex/ | 989511 |
|  | **11** | 9 or 10 | 1385443 |
|  | **12** | 3 and 7 and 8 and 11 | 1307 |
| Web of science  (1985 to 2020)  Indexes=SCI-  EXPANDED, SSCI, A&HCI, CPCI-S, CPCI-SSH, BKCIS, BKCI-SSH, ESCI, CCR-EXPANDED, IC | **1** | TOPIC: (intensive care) OR TOPIC: (critical care) | 204810 |
|  | **2** | TOPIC: (mortality) OR TOPIC: (death) | 1440197 |
|  | **3** | TOPIC: (gender) OR TOPIC: (sex) | 964164 |
|  | **4** | TOPIC: (child*) OR TOPIC: (p*ediatric) | 1508443 |
|  | **5** | #4 AND #3 AND #2 AND #1 | 647 |
|  |  | **Total** | **2411** |

**Table 2. Expanded search updated in December 2020: excluding sex as a search term**

| Database | Search terms | | Number of records |
| --- | --- | --- | --- |
| Ovid MEDLINE^®^  1946 to February Week 4 2020 | **1** | mortality/ or child mortality/ or hospital mortality/ or infant mortality/ or survival rate/ | 282,616 |
|  | **2** | death.mp. or Death/ or Infant Death/ | 824,636 |
|  | **3** | 1 or 2 | 1,041,333 |
|  | **4** | intensive care.mp. | 182,276 |
|  | **5** | critical care.mp. or Critical Care/ | 74,319 |
|  | **6** | exp pediatric intensive care unit/ | 22,778 |
|  | **7** | 4 or 5 or 6 | 221,788 |
|  | **8** | child*.mp. or Child/ | 2,479,668 |
|  | **9** | 3 and 7 and 8 | 5,627 |
| Ovid Embase 1980 to 2020 Week 10 | **1** | mortality/ or child mortality/ or hospital mortality/ or infant mortality/ or survival rate/ | 1,009,665 |
|  | **2** | death.mp. or Death/ or Infant Death/ | 1,270,281 |
|  | **3** | 1 or 2 | 2,060,325 |
|  | **4** | intensive care.mp. | 364,056 |
|  | **5** | critical care.mp. or Critical Care/ | 133,484 |
|  | **6** | exp pediatric intensive care unit/ | 6,857 |
|  | **7** | 4 or 5 or 6 | 377,816 |
|  | **8** | child*.mp. or Child/ | 2,594,888 |
|  | **9** | 3 and 7 and 8 | 15,141 |
| Web of science  (1985 to 2020)  Indexes=SCI-  EXPANDED, SSCI, A&HCI, CPCI-S, CPCI-SSH, BKCIS, BKCI-SSH, ESCI, CCR-EXPANDED, IC | **1** | TOPIC: (intensive care) OR TOPIC: (critical care) | 250,799 |
|  | **2** | TOPIC: (mortality) OR TOPIC: (death) | 1,790,752 |
|  | **3** | TOPIC: (child*) OR TOPIC: (p*ediatric) | 1,944,540 |
|  | **4** | #3 AND #2 AND #1 | 10,854 |
|  |  | **Total** | **31,622** |

**Table 3. Updated search strategy: with expanded search terms for sex**

| Database | Search terms | | Number of records |
| --- | --- | --- | --- |
| Ovid MEDLINE^®^  1946 to December Week 4 2020 | **1** | mortality/ or child mortality/ or hospital mortality/ or infant mortality/ or survival rate/ | 283,324 |
|  | **2** | death.mp. or Death/ or Infant Death/ | 733,560 |
|  | **3** | 1 or 2 | 950,779 |
|  | **4** | intensive care.mp. | 59,093 |
|  | **5** | critical care.mp. or Critical Care/ | 68,566 |
|  | **6** | exp pediatric intensive care unit/ | 22,858 |
|  | **7** | 4 or 5 or 6 | 93,663 |
|  | **8** | child*.mp. or Child/ | 2,316,363 |
|  | **9** | sex.mp. or Sex/ | 785,527 |
|  | **10** | gender.mp. | 291,389 |
|  | **11** | male/ | 8,718,093 |
|  | **12** | female/ | 8,859,939 |
|  | **13** | boy*.mp. | 138,225 |
|  | **14** | girl*.mp. | 133,237 |
|  | **15** | 9 or 10 or 11 or 12 or 13 or 14 | 11,750,175 |
|  | **16** | 3 and 7 and 8 and 15 | 1,992 |
| Ovid Embase  1980 to 2020 Week 52 | **1** | mortality/ or child mortality/ or hospital mortality/ or infant mortality/ or survival rate/ | 1,012,216 |
|  | **2** | death.mp. or Death/ or Infant Death/ | 1,274,634 |
|  | **3** | 1 or 2 | 2,066,744 |
|  | **4** | intensive care.mp. | 708,411 |
|  | **5** | critical care.mp. or Critical Care/ | 57,458 |
|  | **6** | exp pediatric intensive care unit/ | 6,991 |
|  | **7** | 4 or 5 or 6 | 732,165 |
|  | **8** | child*.mp. or Child/ | 2,599,911 |
|  | **9** | sex.mp. or Sex/ | 1,054,497 |
|  | **10** | gender.mp. | 599,318 |
|  | **11** | male/ | 9,224,685 |
|  | **12** | female/ | 9,322,851 |
|  | **13** | boy*.mp. | 206,276 |
|  | **14** | girl*.mp. | 197,109 |
|  | **15** | 9 or 10 or 11 or 12 or 13 or 14 | 12,466,509 |
|  | 16 | 3 and 7 and 8 and 15 | 11,588 |
| Web of science  (2000 to 2020)  Indexes=SCI-  EXPANDED, SSCI, A&HCI, CPCI-S, CPCI-SSH, BKCIS, BKCI-SSH, ESCI, CCR-EXPANDED, IC | 1 | TOPIC: (intensive care) OR TOPIC: (critical care) | 204,810 |
|  | 2 | TOPIC: (mortality) OR TOPIC: (death) | 1,790,752 |
|  | 3 | TOPIC: (child*) OR TOPIC: (p*ediatric) | 1,944,540 |
|  | 4 | TOPIC: (sex) OR TOPIC: (gender) OR TOPIC: (male) OR |  |
|  |  | TOPIC: (female) OR TOPIC: (boy*) OR TOPIC: (girl*) | 2,702,362 |
|  | 5 | #4 AND #3 AND #2 AND #1 | 1,812 |
|  |  | **Total** | **15,392** |
