## Appendix 2 (Tools used in screening, extraction, and quality assessment) for "Is there a sex difference in mortality rates in Paediatric Intensive Care Units: A Systematic Review"

**RefID: 1, There and Back Again: A Review of Residency and Return Migrations in Sharks, with Implications for Population Structure and Management.**
**Chapman DD, Feldheim KA, Papastamatiou Y, Hueter RE**

The overexploitation of sharks has become a global environmental issue in need of a comprehensive and multifaceted management response. Tracking studies are beginning to elucidate how shark movements shape the internal dynamics and structure of populations, which determine the most appropriate scale of these management efforts.

Tracked sharks frequently either remain in a restricted geographic area for an extended period of time (residency) or return to a previously resided-in area after making long-distance movements (site fidelity). Genetic studies have shown that some individuals of certain species preferentially return to their exact birthplaces (natal philopatry) or birth regions (regional philopatry) for either parturition or mating, even though they make long-distance movements that would allow them to breed elsewhere. More than 80 peer-reviewed articles, constituting the majority of published shark tracking and population genetic studies, provide evidence of at least one of these behaviors in a combined 31 shark species from six of the eight extant orders.

Residency, site fidelity, and philopatry can alone or in combination structure many coastal shark populations on finer geographic scales than expected based on their potential for dispersal. This information should therefore be used to scale and inform assessment, management, and conservation activities intended to restore depleted shark populations. Expected final online publication date for the Annual Review of Marine Science Volume 7 is January 03, 2015.

and go to

1. Use the following critera:

   Is this a study in intensive care (if no exclude)

   Are the study participants children (if no exclude)

   Is the study outcome mortality (if no exclude)

   Is mortality grouped by sex (if not sure include)

   Should this study be included

   ◯ yes

   ◯ no

   ◯ maybe

   ◯ keep for introduction

Copyright © 2008-2022, Evidence Partners Inc., All Rights Reserved | Release Notes (https://www.manula.com/manuals/evidence-partners/release-notes/1/en/topic/distillersr)
Loading time: 0.535 seconds

**RefID: 1, There and Back Again: A Review of Residency and Return Migrations in Sharks, with Implications for Population Structure and Management.**
**Chapman DD, Feldheim KA, Papastamatiou Y, Hueter RE**

The overexploitation of sharks has become a global environmental issue in need of a comprehensive and multifaceted management response. Tracking studies are beginning to elucidate how shark movements shape the internal dynamics and structure of populations, which determine the most appropriate scale of these management efforts.

Tracked sharks frequently either remain in a restricted geographic area for an extended period of time (residency) or return to a previously resided-in area after making long-distance movements (site fidelity). Genetic studies have shown that some individuals of certain species preferentially return to their exact birthplaces (natal philopatry) or birth regions (regional philopatry) for either parturition or mating, even though they make long-distance movements that would allow them to breed elsewhere. More than 80 peer-reviewed articles, constituting the majority of published shark tracking and population genetic studies, provide evidence of at least one of these behaviors in a combined 31 shark species from six of the eight extant orders.

Residency, site fidelity, and philopatry can alone or in combination structure many coastal shark populations on finer geographic scales than expected based on their potential for dispersal. This information should therefore be used to scale and inform assessment, management, and conservation activities intended to restore depleted shark populations. Expected final online publication date for the Annual Review of Marine Science Volume 7 is January 03, 2015.

and go to

1. Use the following critera then answer the question

   Screen tehe full text article for the following:

   - Is the population children (if no exclude)
   - Is the population mixed adults and children (if yes exclude)
   - Is the setting intensive care (if no exclude)
   - Is mortality listed as an outcome (if no exclude)
   - Is mortality grouped by sex (if no exclude)

   Should this study be included

   ◯ yes

   ◯ no

2. What is the reason for exclusion

   ☐ Mortality is not an outcome

   ☐ Mortality is outside PICU

   ☐ Population is not children

   ☐ Mixed adults and children

   ☐ Setting is not PICU

   ☐ Mortality is not reported by sex

   ☐ Included

   ☐ Full text not available in English

   ☐ Abstract only - no full text

   ☐ Mortality is part of a composite outcome

   ☐ Unable to extract data

   ☐ No observed mortality

   ☐ Case series

   ☐ Neonates only

   ☐ Protocol only

   ☐ Full text not available

   ☐ Other

   **Permanently add an answer to this question ()**

3. Is sex the primary exposure

   ◯ Yes

   ◯ No

   ◯ NA

Copyright © 2008-2022, Evidence Partners Inc., All Rights Reserved | Release Notes (https://www.manula.com/manuals/evidence-partners/release-notes/1/en/topic/distillersr)
Loading time: 0.305 seconds

### Applying inclusion and exclusion criteria

**RefID: 1, There and Back Again: A Review of Residency and Return Migrations in Sharks, with Implications for Population Structure and Management.**
**Chapman DD, Feldheim KA, Papastamatiou Y, Hueter RE**


The overexploitation of sharks has become a global environmental issue in need of a comprehensive and multifaceted management response. Tracking studies are beginning to elucidate how shark movements shape the internal dynamics and structure of populations, which determine the most appropriate scale of these management efforts.

Tracked sharks frequently either remain in a restricted geographic area for an extended period of time (residency) or return to a previously resided-in area after making long-distance movements (site fidelity). Genetic studies have shown that some individuals of certain species preferentially return to their exact birthplaces (natal philopatry) or birth regions (regional philopatry) for either parturition or mating, even though they make long-distance movements that would allow them to breed elsewhere. More than 80 peer-reviewed articles, constituting the majority of published shark tracking and population genetic studies, provide evidence of at least one of these behaviors in a combined 31 shark species from six of the eight extant orders.

Residency, site fidelity, and philopatry can alone or in combination structure many coastal shark populations on finer geographic scales than expected based on their potential for dispersal. This information should therefore be used to scale and inform assessment, management, and conservation activities intended to restore depleted shark populations. Expected final online publication date for the Annual Review of Marine Science Volume 7 is January 03, 2015.


and go to

1. Reviewer name

...

### Study characteristics & eligibility

2. Study Design

Study design is used to determine which Risk of Bias Assessment type is used. RCTs, Controlled Trials, Controlled Before and After trials, and N-of-1 trials are assessed using the Cochrane Risk of Bias Tool. Cohort and case control studies are assessed using the Newcastle Ottawa Scale. Other study designs are generally considered to be at high risk of bias and so a simple summary form is used to note exceptional factors. You may develop additional risk of bias forms for particular designs.

Select an Answer

**Add a study design ()**

3. Setting

- ☐ PICU
- ☐ ICU
- ☐ Neonatal ICU
- ☐ Unclear
- ☐ Cardiac ICU
- ☐ Cardiac PICU

**Add an institution type ()**

4. Whole PICU poplulation or subgroup

- ◯ Whole PICU
- ◯ Subgroup

### Participants

6. Age range of participants

...

### Types of outcome measures

7. Mortality outcome

Tick all that apply

- ☐ Outcome is death in PICU
- ☐ 7 day mortality
- ☐ 14 day mortality

- [ ] 30 day mortality
- [ ] Death within 48 hours of PICU admission
- [ ] Unclear - refers to
- [ ] 3 day mortality
- [ ] Overall mortality
- [ ] In hospital mortality
- [ ] No mortality outcome given
- [ ] 60 day mortality
- [ ] 90 day mortality
- [ ] 28 day mortality
- [ ] Death within 24 hours of decannulation
- [ ] Death within 24 hours of PICU admission
- [ ] Death within 1 year of PICU admission
- [ ] 5 year mortality
- [ ] Death 2 months after admission
- [ ] Death 28 days after PICU discharge
- [ ] Death 6 months after PICU discharge
- [ ] Death after PICU discharge
- [ ] death 30 days after PICU discharge
- [ ] 6 month mortality
- [ ] Mortlaity in first 96h of hospital admission
- [ ] 2 year mortality

**Add a different mortality outcome ()**

8. Exposure (sex)

- [ ] Sex was the primary exposure
- [ ] Mortality grouped by sex
- [ ] Invasive procedures
- [ ] CI measurement
- [ ] Chronic Conditions
- [ ] Mode of renal replacement therapy
- [ ] Serum uric acid
- [ ] Chronic health conditions

**Other exposure grouping ()**

### Study eligibility

Exclude at this stage before any data exctraction takes place

9. Should this study be included

◯ Yes

◯ No

**Clear Response ()**

11. Reason for exclusion

- ◯ Cannot retrieve full report
- ◯ Data in format that cannot be extracted
- ◯ N/A
- ◯ Does not seperate PICU mortality from overall mortality
- ◯ No mortality outcome
- ◯ Excludes death on first day of PICU admission
- ◯ NICU study
- ◯ Protocol only, no results
- ◯ Does not seperate adult from paediatric patients
- ◯ Unclear if mortality is in PICU
- ◯ Mortlaity not reported by sex
- ◯ Unclear age range
- ◯ Case series
- ◯ Excludes admissions <48h
- ◯ Excludes admissions <72h
- ◯ Excludes admissions <24h
- ◯ Mortality within 48h only
- ◯ Reported elsewhwre

**Add other reasons ()**

Copyright © 2008-2022, Evidence Partners Inc., All Rights Reserved | Release Notes (https://www.manula.com/manuals/evidence-partners/release-notes/1/en/topic/distillersr)
Loading time: 0.364 seconds

Data extraction

**RefID: 1, There and Back Again: A Review of Residency and Return Migrations in Sharks, with Implications for Population Structure and Management. Chapman DD, Feldheim KA, Papastamatiou Y, Hueter RE**

The overexploitation of sharks has become a global environmental issue in need of a comprehensive and multifaceted management response. Tracking studies are beginning to elucidate how shark movements shape the internal dynamics and structure of populations, which determine the most appropriate scale of these management efforts.

Tracked sharks frequently either remain in a restricted geographic area for an extended period of time (residency) or return to a previously resided-in area after making long-distance movements (site fidelity). Genetic studies have shown that some individuals of certain species preferentially return to their exact birthplaces (natal philopatry) or birth regions (regional philopatry) for either parturition or mating, even though they make long-distance movements that would allow them to breed elsewhere. More than 80 peer-reviewed articles, constituting the majority of published shark tracking and population genetic studies, provide evidence of at least one of these behaviors in a combined 31 shark species from six of the eight extant orders.

Residency, site fidelity, and philopatry can alone or in combination structure many coastal shark populations on finer geographic scales than expected based on their potential for dispersal. This information should therefore be used to scale and inform assessment, management, and conservation activities intended to restore depleted shark populations. Expected final online publication date for the Annual Review of Marine Science Volume 7 is January 03, 2015.

and go to

### Participants

1. Number of males

...

2. Number of females

...

3. Total number of participants

...

4. % of male admissions

...

5. % of female admissions

...

6. Subgroup

**Clear Response ()**

### Risk Ratio (RR), Odds Ratio (OR), Relative Risk Difference (RD) and Confidence Intervals (CI)

This form calculates crude relative risk using the following formula:

a = No. deaths in females

b = Total no. females

c = No. deaths in males

d = Total no. males

Relative Risk = (a/b)/(c/d)

Odd Ratio = (a*(d-c))/((b-c)*c)

Risk Difference = (a/b)-(c/d)

RR = (a/b) / (c/d)

| Number of PICU **Deaths** in **Females** | Number of **Females** in PICU |
|---|---|
| ... | ... |
| Number of PICU **Deaths** in **Males** | Number of **Males** in PICU |
| ... | ... |

### Risk and Odds Ratios

Female relative to Male

| Risk Ratio (Calculated) | OR (Calculated) |
| --- | --- |
| ... | ... |
| ln(OR) (Calculated) | SE ln(OR) (Calculated) |
| -Infinity | Infinity |

#### Confidence Interval

| LL log OR (Calculated) | UL log OR (Calculated) |
| --- | --- |
| -Infinity | ... |
| LL 95% CI OR (Calculated) | UL 95% CI OR (Calculated) |
| 0 | 1 |

#### Risk Difference (Automatically Calculated)

Female - Male

...

#### Relative Risk

Female relative to Male

| RR (Calculated) | |
| --- | --- |
| ... | |
| ln(RR) (Calculated) | SE ln(RR) (Calculated) |
| -Infinity | ... |
| LL 95% CI RR (Calculated) | UL 95% CI RR (Calculated) |
| -Infinity | -Infinity |

#### Adjusted estimate

If available

25. Add the adjusted/reported estimate

...

26. Confidence interval

...

27. Other adjustment variables

- [ ] Age
- [ ] PIM
- [ ] Race/Ethnicity
- [ ] Insurance type
- [ ] Primary diagnosis
- [ ] Time period
- [ ] High risk conditions
- [ ] SES
- [ ] Serum Chloride
- [ ] Admission time
- [ ] Origin of admission
- [ ] Centre/Unit
- [ ] Staffing
- [ ] Trauma
- [ ] PRISM
- [ ] Previous ICU/PICU
- [ ] Inotropes
- [ ] Post-op care
- [ ] Chronic Conditions
- [ ] TBI
- [ ] qSOFA
- [ ] Sepsis
- [ ] Mechanical ventilation
- [ ] Malaria parasite
- [ ] Hyperglycemia
- [ ] Valiables not reported
- [ ] No. organ dysfunction
- [ ] PLOD
- [ ] Unadjusted
- [ ] LOS
- [ ] Planned/unplanned admission
- [ ] Days on ECMO
- [ ] Mode of ECMO
- [ ] Wt change
- [ ] Nitric Oxide
- [ ] Renal replacement
- [ ] Congenital heart disease
- [ ] Blood oxygen
- [ ] Out-of-hospital arrest

- [ ] Transfer to PICU
- [ ] Neurological disorders
- [ ] Prior Neurodevelopment
- [ ] EEG Background Category
- [ ] Seizure Category
- [ ] GCS
- [ ] Steroids
- [ ] Nutritional status
- [ ] Antibiotics

**Add an adjustment variable ()**

### Length of stay and comments

28. LOS

    Enter the LOS for females/male. See examples below:

    - Median days 3/3.5
    - Mean days <4/<4
    - No difference
    - Not reported

    ...

29. Additional comments if any

    ...

30. Reviewer name

    ...

Copyright © 2008-2022, Evidence Partners Inc., All Rights Reserved | Release Notes (https://www.manula.com/manuals/evidence-partners/release-notes/1/en/topic/distillersr)
Loading time: 0.369 seconds

### Extended extraction for studies where sex is the main exposure

**RefID: 1, There and Back Again: A Review of Residency and Return Migrations in Sharks, with Implications for Population Structure and Management.**
**Chapman DD, Feldheim KA, Papastamatiou Y, Hueter RE**

The overexploitation of sharks has become a global environmental issue in need of a comprehensive and multifaceted management response. Tracking studies are beginning to elucidate how shark movements shape the internal dynamics and structure of populations, which determine the most appropriate scale of these management efforts.

Tracked sharks frequently either remain in a restricted geographic area for an extended period of time (residency) or return to a previously resided-in area after making long-distance movements (site fidelity). Genetic studies have shown that some individuals of certain species preferentially return to their exact birthplaces (natal philopatry) or birth regions (regional philopatry) for either parturition or mating, even though they make long-distance movements that would allow them to breed elsewhere. More than 80 peer-reviewed articles, constituting the majority of published shark tracking and population genetic studies, provide evidence of at least one of these behaviors in a combined 31 shark species from six of the eight extant orders.

Residency, site fidelity, and philopatry can alone or in combination structure many coastal shark populations on finer geographic scales than expected based on their potential for dispersal. This information should therefore be used to scale and inform assessment, management, and conservation activities intended to restore depleted shark populations. Expected final online publication date for the Annual Review of Marine Science Volume 7 is January 03, 2015.

and go to

1. Reviewer name

...

#### Study characteristics

2. Whole PICU population or subgroup

◯ Whole population
◯ Sepsis
◯ Malignancy
◯ Cardiac
◯ Burns
◯ Patients with Diarrhoea
◯ Severe Health Conditions
◯ Life-threatening influenza infection

**Add population type eg sepsis ()**

**Clear Response ()**

3. Date range of the study

...

#### Geographic Locations

Instructions to review coordinator. Edit initial locations lists as appropriate.

4. Geographical location, principal site:

...

5. Number of sites

...

6. Clusters

eg Hospitals, ICUs, regions etc

☐ ICUs/PICUs
☐ Hospitals
☐ Regions
☐ Single centre

**Permanently add an answer to this question ()**

#### Participants

7. Number of females

...

8. Number of males

...

9. Total number of participants

...

10. % of male admissions

...

11. % of female admissions

...

12. Age range of participants

48 hours - 18 years

13. Population description

If there are inclusion and exclusion criteria that are not already collected in this form add them here

...

14. Method of recruitment

...

15. Baseline imbalances if any

...

16. Race/Ethnicity

...

17. Severity of illness

What score was used

- [ ] PIM
- [ ] PRISM
- [ ] None

**Add any other score ()**

18. Comorbidities

Please list if relevant or reported. Otherwise please state none reported or any appropriate response

...

19. Other relevant demographics

If none please state "none"

...

20. Subgroup measures

Add subgroups as needed

**Add a subgroup ()**

21. Any relevant notes

...

22. Exclude?

◯ Yes

◯ No

**Clear Response ()**

### Outcome

24. Mortality is the primary outcome

◯ Yes

◯ No

**Clear Response ()**

25. Mortality outcome

Tick all that apply

☐ Outcome is death in PICU

☐ 7 day mortality

☐ 14 day mortality

☐ 30 day mortality

**Add a different mortality outcome ()**

26. Exposure (sex)

☐ Sex was the primary exposure

☐ Mortality grouped by sex

**Other exposure grouping ()**

### Study eligibility

Exclude at this stage before any data exctraction takes place

27. Suitable for inclusion in Meta-Analysis?

◯ Yes

◯ No

**Clear Response ()**

### Calculations

This form calculates crude relative risk using the following formula:

a = No. deaths in females

b = Total no. females

c = No. deaths in males

d = Total no. males

Relative Risk = (a/b)/(c/d)

Odd Ratio = (a*(d-c))/((b-c)*c)

Risk Difference = (a/b)-(c/d)

RR = (a/b) / (c/d)

Number of PICU **Deaths** in **Females**

...

Number of **Females** in PICU

...

Number of PICU **Deaths** in **Males**

...

Number of **Males** in PICU

...

### Risk and Odds Ratios

Female relative to Male

| | |
|---|---|
| Risk Ratio (Automatically Calculated)<br>... | Odds Ratio (Automatically Calculated)<br>... |
| ln(OR) (Automatically Calculated)<br>-Infinity | SE ln(OR) (Automatically Calculated)<br>Infinity |

### Confidence Interval

| | |
|---|---|
| Lower Limit of OR Confidence Interval (Automatically Calculated)<br>-Infinity | Upper Limit of OR Confidence Interval (Automatically Calculated)<br>... |
| Lower Limit of the 95% CI of the OR (Automatically Calculated)<br>0 | Upper Limit of the 95% CI of the OR (Automatically Calculated)<br>1 |

### Risk Difference (Automatically Calculated)

Female - Male

...

### Relative Risk

Female relative to Male

| | |
|---|---|
| Relative Risk (Automatically Calculated)<br>... | |
| ln(Relative Risk) (Automatically Calculated)<br>-Infinity | SE ln(Relative Risk) (Automatically Calculated)<br>... |

| Lower Limit of the 95% CI of Relative Risk (Automatically Calculated) | Upper Limit of the 95% CI of Relative Risk (Automatically Calculated) |
|---|---|
| -Infinity | -Infinity |

For the questions below, add answere if provided in the paper

46. Calculated estimates such as adjusted or unadjusted OR, RR

◯ Provided

◯ Not provided

**Clear Response ()**

48. What is the value of the estimate

...

49. What is the baseline group

◯ Males/Female

◯ Females/Males

◯ Males - Females

◯ Females - Males

**Add another ()**

**Clear Response ()**

50. What are the confidence intervals

...

51. If the estimate is adjusted, list the variables

☐ No adjustment

☐ Age

☐ Admission diagnosis

☐ Nosocomial infection

☐ Ethnicity

☐ Season

☐ Deprivation score

☐ PIM2

☐ PICU

**Add an adjustment variable ()**

**Copyright © 2008-2022, Evidence Partners Inc., All Rights Reserved | Release Notes (https://www.manula.com/manuals/evidence-partners/release-notes/1/en/topic/distillersr)**
**Loading time: 0.336 seconds**

### Modified ROBINS-E for quality assessment of studies where sex is the main exposure

**RefID: 1, There and Back Again: A Review of Residency and Return Migrations in Sharks, with Implications for Population Structure and Management.**
**Chapman DD, Feldheim KA, Papastamatiou Y, Hueter RE**


The overexploitation of sharks has become a global environmental issue in need of a comprehensive and multifaceted management response. Tracking studies are beginning to elucidate how shark movements shape the internal dynamics and structure of populations, which determine the most appropriate scale of these management efforts.

Tracked sharks frequently either remain in a restricted geographic area for an extended period of time (residency) or return to a previously resided-in area after making long-distance movements (site fidelity). Genetic studies have shown that some individuals of certain species preferentially return to their exact birthplaces (natal philopatry) or birth regions (regional philopatry) for either parturition or mating, even though they make long-distance movements that would allow them to breed elsewhere. More than 80 peer-reviewed articles, constituting the majority of published shark tracking and population genetic studies, provide evidence of at least one of these behaviors in a combined 31 shark species from six of the eight extant orders.

Residency, site fidelity, and philopatry can alone or in combination structure many coastal shark populations on finer geographic scales than expected based on their potential for dispersal. This information should therefore be used to scale and inform assessment, management, and conservation activities intended to restore depleted shark populations. Expected final online publication date for the Annual Review of Marine Science Volume 7 is January 03, 2015.


and go to

#### Risk of Bias in Cohort Studies

**Country**

Select an Answer

2. Was selection of exposed and non-exposed cohorts drawn from the same population?

Definitely yes (low risk of bias)

Probably yes

Probably no

Definitely no (high risk of bias)

- **Examples of low risk of bias:**
  - Exposed and unexposed drawn for same administrative data base of patients presenting at same points of care over the same time frame
- **Examples of high risk of bias:**
  - Exposed and unexposed presenting to different points of care or over a different time frame

3. Can we be confident in the assessment of exposure?

Definitely yes (low risk of bias)

Probably yes

Probably no

Definitely no (high risk of bias)

- **Examples of low risk of bias:**
  - Secure record (e.g. surgical records, pharmacy records)
  - Repeated interview or other ascertainment asking about current use/exposure
- **Examples of higher risk of bias:**
  - Structured interview at a single point in time
  - Written self report
  - Individuals who are asked to retrospectively confirm their exposure status may be subject to recall bias – less likely to recall an exposure if they have not developed an adverse outcome, and more likely to recall an exposure (whether an exposure occurred or not) if they have developed an adverse outcome
- **Examples of high risk of bias:**
  - Uncertain how exposure information obtained

4. Can we be confident that the outcome of interest was not present at start of study?

Definitely yes (low risk of bias)

Probably yes

Probably no

Definitely no (high risk of bias)

5. Did the study match exposed and unexposed for all variables that are associated with the outcome of interest or did the statistical analysis adjust for these prognostic variables?

Definitely yes (low risk of bias)

Mostly yes

Mostly no

Definitely no (high risk of bias)

- **Examples of low risk of bias:**
    - Comprehensive matching or adjustment for all plausible prognostic variables
- **Examples of higher risk of bias:**
    - Matching or adjustment for most plausible prognostic variables
- **Examples of high risk of bias:**
    - Matching or adjustment for a minority of plausible prognostic variables, or no matching or adjustment at all
    - Statements of no differences between groups or that differences were not statistically significant are not sufficient for establishing comparability

6. Can we be confident in the assessment of the presence or absence of prognostic factors?

Definitely yes (low risk of bias)

Probably yes

Probably no

Definitely no (high risk of bias)

- **Examples of low risk of bias:**
    - Interview of all participants
    - Self-completed survey from all participants
    - Review of charts with reproducibility demonstrated
    - From data base with documentation of accuracy of abstraction of prognostic data
- **Examples of higher risk of bias:**
    - Chart review without demonstration of reproducibility
    - Data base with uncertain quality of abstraction of prognostic information
- **Examples of high risk of bias:**
    - Prognostic information from data base with no available documentation of quality of abstraction of prognostic variables

7. Can we be confident in the assessment of outcome?

Definitely yes (low risk of bias)

Probably yes

Probably no

Definitely no (high risk of bias)

- **Examples of low risk of bias:**
    - Independent blind assessment
    - Record linkage
    - For some outcomes (e.g. fractured hip), reference to the medical record is sufficient to satisfy the requirement for confirmation of the fracture
- **Examples of higher risk of bias:**
    - Independent assessment unblinded
    - Self-report
    - For some outcomes (e.g. vertebral fracture where reference to x-rays would be required) reference to the medical record would not be adequate outcomes
- **Examples of high risk of bias:**
    - Uncertain (no description)

8. Was the follow up of cohorts adequate?

- Definitely yes (low risk of bias)
- Probably yes
- Probably no
- Definitely no (high risk of bias)

- **Examples of low risk of bias:**
    - No missing outcome data
    - Reasons for missing outcome data unlikely to be related to true outcome (for survival data, censoring is unlikely to introduce bias)
    - Missing outcome data balanced in numbers across intervention groups, with similar reasons for missing data across groups
    - For dichotomous outcome data, the proportion of missing outcomes compared with observed event risk is not enough to have a important impact on the intervention effect estimate
    - For continuous outcome data, plausible effect size (difference in means or standardized difference in means) among missing outcomes is not large enough to have an important impact on the observed effect size
    - Missing data have been imputed using appropriate methods
- **Examples of high risk of bias:**
    - Reason for missing outcome data likely to be related to true outcome, with either imbalance in numbers or reasons for missing data across intervention groups
    - For dichotomous outcome data, the proportion of missing outcomes compared with observed event risk is enough to induce important bias in intervention effect estimate
    - For continuous outcome data, plausible effect size (difference in means or standardized difference in means) among missing outcomes is large enough to induce clinically relevant bias in the observed effect size

9. Were co-Interventions similar between groups?

- Definitely yes (low risk of bias)
- Probably yes
- Probably no
- Definitely no (high risk of bias)

- **Examples of low risk of bias:**
    - Most or all relevant co-interventions that might influence the outcome of interest are documented to be similar in the exposed and unexposed
- **Examples of high risk of bias:**
    - Few or no relevant co-interventions that might influence the outcome of interest are documented to be similar in the exposed and unexposed

### Assessment of Bias

(*Automatically Generated*)

- Low risk of bias for all key domains.
- Unclear risk of bias for one or more key domains.
- High risk of bias for one or more key domains.

**Clear Response ()**

##### Citing this tool


Busse JW, Guyatt GH. Tool to Assess Risk of Bias in Cohort Studies. https://www.evidencepartners.com/resources/methodological-resources/

Busse JW, Guyatt GH. Tool to Assess Risk of Bias in Case-control Studies. https://www.evidencepartners.com/resources/methodological-resources/

Guyatt GH, Busse JW. Modification of Cochrane Tool to assess risk of bias in randomized trials. https://www.evidencepartners.com/resources/methodological-resources/

Tikkinen K, Busse J, Guyatt G. Tool to assess risk of bias in observational studies of natural history of medical symptoms/conditions in general populations. https://www.evidencepartners.com/resources/methodological-resources/


EP 2020


**Copyright © 2008-2022, Evidence Partners Inc., All Rights Reserved | Release Notes (https://www.manula.com/manuals/evidence-partners/release-notes/1/en/topic/distillersr)**

**Loading time: 0.311 seconds**
