## Appendix 3. Tables of study summaries for "Is there a sex difference in mortality rates in Paediatric Intensive Care Units: A Systematic Review"

### Table1. Summary of studies reporting higher female mortality

| Study | Design | Age | Population | % Male | F/M OR | Reported | Estimate | Adjustment |
|---|---|---|---|---|---|---|---|---|
| Shanmugham 2018[1] | Prospective | 0 - 12y | MV | 54.62 | 10.76 | | | |
| Rowan 2016[2] | Prospective | median 8.7y | BMT | 56.31 | 5.06 | | | |
| Ali 2016[3] | Retrospective | 0 - 18y | Oncology | 60 | 4.9 | | | |
| Thukral 2006[4] | Prospective | Median 18m | Whole PICU | 63.26 | 3.73 | | | |
| Coetzee 2014[5] | Retrospective | 0 - 7y | Measles | 44.83 | 2.87 | | | |
| Sayed 2018[6] | Prospective | 1 - 15y | Oncology | 63.41 | 2.64 | | | |
| Patel 2017[7] | Retrospective | 0 - 15y | Trauma | 57.72 | 2.61 | | | |
| Selewski 2012[8] | Retrospective | 0 - 10m | ECMO | 60.38 | 2.19 | | | |
| Lee 2017[9] | Retrospective | 0 - 18y | Abuse/Maltreatment | 65.35 | 2.17 | | | |
| Aroor 2018[10] | Prospective | 0 - 18y | Whole PICU | 62.31 | 2.15 | | | |
| Topjian 2013[11] | Prospective | >1m | Seizures | 56 | 2.03 | OR f/m | 2.5 | Age |
| Jacobe 2003[12] | Retrospective | <3.81y | BMT | 72.5 | 2.02 | | | |
| Hui 2012[13] | Retrospective | 0 - 18y | RRT | 59.46 | 2 | | | |
| Hardelid 2018[14] | Retrospective | <16y | Whole PICU | 57.73 | 1.99 | OR f/m | 1.91 | Age, Race/Ethnicity, Time period, High risk conditions, SES |
| Al-Ayed 2018[15] | Prospective | 0 - 14y | RRT | 56.25 | 1.98 | | | |

| Study | Design | Age | Population | % Male | F/M OR | Reported | Estimate | Adjustment |
|---|---|---|---|---|---|---|---|---|
| Parajuli 2020[16] | Retrospective | 0 - 12y | Hemophagocytic Lymphohistiocytosis | 59.68 | 1.81 | | | |
| Koh 2017[17] | Retrospective | <18y | Pneumonia/Resp tract infection | 53.16 | 1.8 | | | |
| Sinitsky 2015[18] | Retrospective | 0 - 16y | MV | 56.67 | 1.79 | | | |
| de Souza 2016[19] | Prospective | 0 - 17y | Sepsis or shock | 52.41 | 1.79 | OR f/m | 2.25 | Variables not reported |
| Jeschke 2008[20] | Retrospective | 0 - 16y | Burns | 59.79 | 1.78 | | | |
| Hariharan 2011[21] | Prospective | median 5y | Whole PICU | 57.67 | 1.76 | | | |
| Paret 1999[22] | Retrospective | 50d - 16y | ARDS | 61.54 | 1.75 | | | |
| Bekhit 2014[23] | Retrospective | 0 - 168m | Whole PICU | 51.22 | 1.73 | | | |
| Mitra 2000[24] | Retrospective | <5y | Patients with Diarrhoea | 63.33 | 1.65 | | | |
| Egbohou 2019[25] | Retrospective | 0 - 15y | Whole PICU | 64.23 | 1.64 | | | |
| Berndtson 2013[26] | Prospective | <18y | Burns | 62.65 | 1.6 | | | |
| Erdem 2019[27] | Prospective | <18y | ECMO | 52.94 | 1.57 | | | |
| Li 2015[28] | Retrospective | median 11/12m | Whole PICU | 63.31 | 1.52 | | | |
| Abebe 2015[29] | Cross-sectional | 0 - 14y | Whole PICU | 54.71 | 1.52 | | | |
| Malhotra 2020[30] | Retrospective | 0 - 15y | Whole PICU | 61.62 | 1.52 | | | |
| Esteban 2015[31] | Retrospective | 0 - 18+y | Severe Health Conditions | 57.26 | 1.5 | | | |
| An 2016[32] | Retrospective | median 6y | BMT | 68.97 | 1.5 | | | |

| Study | Design | Age | Population | % Male | F/M OR | Reported | Estimate | Adjustment |
|---|---|---|---|---|---|---|---|---|
| Khemani 2009[33] | Retrospective | <18y | Sepsis or shock | 62.88 | 1.49 | | | |
| Odetola 2005[34] | Retrospective | median 19m | Meningitis | 57.42 | 1.46 | | | |
| Earan 2016[35] | Retrospective | 0 - 15y | Whole PICU | 63.61 | 1.41 | | | |
| Egbohou 2019[36] | Retrospective | 0 - 15y | TBI/Head injury | 67.03 | 1.38 | | | |
| Hsiao 2015[37] | Retrospective | 0 - 12y | Pneumococcal disease | 39.58 | 1.33 | | | |
| Scholefield 2015[38] | Retrospective | 0 - 16y | Out of hospital cardiac arrest | 58.4 | 1.33 | | | |
| Ping 2018[39] | Prospective | median 1.4y | Long stay | 51.45 | 1.32 | | | |
| Verlaat 2019[40] | Retrospective | <18y | Complex Chronic Conditions | 57.72 | 1.3 | OR m/f | 0.75 | Age, Primary diagnosis, Admission time, Chronic Conditions |
| Bhaskar 2015[41] | Case | 0 - 18y | Sepsis or shock | 58.77 | 1.29 | | | |
| Choudhary 2020[42] | Prospective | 0 - 14y | RRT | 62 | 1.27 | | | |
| Du 2020[43] | Retrospective | <16y | Whole PICU | 58.29 | 1.27 | | | |
| Riyuzo 2017[44] | Retrospective | 1 - 12y | AKI | 61.04 | 1.24 | | | |
| Punchak 2018[45] | Retrospective | 0 - 15y | Whole PICU | 56.82 | 1.21 | | | |
| Iro 2019[46] | Retrospective | 0 - 17y | Encephalitis | 55 | 1.21 | | | |
| Balit 2016[47] | Retrospective | <18y | BMT | 59.22 | 1.2 | OR f/m | 1.2 | Unadjusted |
| Sachdev 2018[48] | Prospective | median 72m | Whole PICU | 67.33 | 1.2 | | | |

| Study | Design | Age | Population | % Male | F/M OR | Reported | Estimate | Adjustment |
|---|---|---|---|---|---|---|---|---|
| Mahdi 2018[49] | Cross-sectional | <15y | Whole PICU | 59.09 | 1.19 | | | |
| Krishnamurthy 2013[50] | Prospective | 0 - 13y | AKI | 53.7 | 1.14 | | | |
| Patki 2014[51] | Prospective | 0 - 16y | Whole PICU | 64.36 | 1.14 | | | |
| Ferreira 2020[52] | Retrospective | 0 - 13y | AKI | 56.68 | 1.14 | | | |
| Polito 2020[53] | Retrospective | <16y | Whole PICU | 57.16 | 1.14 | | | |
| Horoz 2019[54] | Prospective | 0 - 18y | Intra-abdominal hypertension | 54.29 | 1.11 | | | |
| Raymakers-Janssen 2019[55] | Retrospective | median 8.9y | BMT | 63.24 | 1.11 | | | |
| Fraser 2018[56] | Retrospective | 0 - 16y | Life-limiting conditions | 56.71 | 1.09 | OR f/m | 1.09 | Age, PIM, Race/Ethnicity, Primary diagnosis, SES, Centre/Unit |
| Verlaat 2017[57] | Prospective | <18y | Low risk PICU patients | 57.5 | 1.07 | | | |
| Yadav 2019[58] | Prospective | 0 - 14y | ARDS | 66.94 | 1.07 | | | |
| Epstein 2011[59] | Retrospective | 0 - 18y | Whole PICU | 55.8 | 1.06 | OR f/m | 1.12 | Age, PIM, Race/Ethnicity, Insurance type, Primary diagnosis |
| AbdAllah 2016[60] | Observation | 0 - 15y | Whole PICU | 51.2 | 1.06 | | | |
| AlKadhem 2020[61] | Prospective | 0 - 14y | Whole PICU | 53.24 | 1.04 | | | |
| Klein 2008[62] | Prospective | Mean 82.4m | Whole PICU | 51.94 | 1.03 | | | |

| Study | Design | Age | Population | % Male | F/M OR | Reported | Estimate | Adjustment |
|---|---|---|---|---|---|---|---|---|
| McCrory 2017[63] | Retrospective | <18y | Whole PICU | 56 | 1.03 | OR f/m | 1.03 | Age, PIM, Admission time, Origin of admission, Centre/Unit, Staffing, Trauma |
| Moynihan 2019[64] | Retrospective | <16y | Whole PICU | 57.26 | 1.02 | | | |
| Alobaidi 2020[65] | Retrospective | 0 - 17y | AKI | 56.54 | 1.01 | | | |
| Branco 2005[66] | Prospective | mean 33.8m | Sepsis or shock | 66.67 | | OR m/f | 0.7 | PRISM, Hyperglycaemia |
| Jaramillo-Bustamante 2012[67] | Prospective | 0 - 18y | Sepsis or shock | 55 | | % m/f | 18% vs 18.5% | Unadjusted |
| Ferguson 2012[68] | Prospective | 0 - 16y | Arrest | 57.92 | | OR m/f | 0.86 | Age, PIM, Race/Ethnicity, Time period, Inotropes, Mechanical ventilation |

### Table 2. Summary of studies reporting equal male and female mortality

| Study | Design | Age | Population | % Male | F/M OR | Reported | Estimate | Adjustment |
|---|---|---|---|---|---|---|---|---|
| Cairo 2018[69] | Prospective | 0 - 18y | ECMO | 53.14 | | OR m/f dx alive | 0.98 | Age, Race/Ethnicity, Time period, Mechanical ventilation |
| Tyagi 2018[70] | Prospective | 0 - 12y | Whole PICU | 56.29 | 0.98 | | | |
| Bejersten 1988[71] | Prospective | 0 - 15y | Whole PICU | 56.05 | 0.99 | | | |
| Volakli 2012[72] | Prospective | 0 - 14y | Whole PICU | 65.33 | 0.99 | | | |
| Purcell 2020[73] | Prospective | 0 - 18y | Whole PICU | 43.81 | 1 | RR f/m | 0.97 | Age, Inotropes, TBI, qSOFA, Sepsis, Mechanical ventilation, Malaria parasite |
| Arias 2004[74] | Retrospective | Children | Whole PICU | 57.27 | | OR m/f | 1 | Age, Admission time, Origin of admission, PRISM, Previous ICU/PICU, Inotropes, Post-op care |

F/M OR: This is the female/male OR we calculated from the crude numbers provided, Reported: This is the measure provided by the authors of the study, Adjustment: Any variables used for adjustment for the reported measure, AKI: Acute kidney injury, ARDS: Acute respiratory distress syndrome, Arrest: Cardiorespiratory arrest, BG: Blood glucose, ECMO: Extra-corporeal membrane oxygenation, MOF: Multi organ failure, MV: Mechanical ventilation, RRT: Renal replacement therapy, TBI: Traumatic brain injury, SES: Socio-economic status

**Table 3. Summary of studies reporting higher male mortality**

| Study | Design | Age | Population | % Male | F/M OR | Reported | Estimate | Adjustment |
|---|---|---|---|---|---|---|---|---|
| Hon 2016[75] | Retrospective | <12y | Encephalitis | 60.87 | 0.14 | | | |
| Dursun 2020[76] | Retrospective | <18y | Oncology | 43.75 | 0.18 | | | |
| Wang 2008[77] | Prospective | 0 - 18y | HFOV | 60.61 | 0.19 | | | |
| Piastra 2019[78] | Retrospective | <14y | Cerebral haemorrhage | 56.07 | 0.29 | | | |
| Zobel 1991[79] | Retrospective | mean 4.3y | RRT | 69.23 | 0.36 | | | |
| Branco 2011[80] | Prospective | 0 - 16y | MV | 47.92 | 0.4 | | | |
| Berger 2013[81] | Prospective | <18y | Pertussis infection | 51.15 | 0.49 | | | |
| Lefevre 2017[82] | Retrospective | 0 - 12y | Sepsis or shock | 53.52 | 0.51 | | | |
| Lombel 2012[83] | Retrospective | median 184m | BMT | 66.67 | 0.53 | | | |
| Torres 2012[84] | Retrospective | 0 - 110m | Influenza | 60.56 | 0.56 | | | |
| Nyirasafari 2017[85] | Prospective | 0 - 15y | Whole PICU | 60 | 0.57 | OR f/m | 0.63 | Nutritional status, diagnosis, modified PRISM score, CPAP, vaso-active drugs |
| Kaur 2014[86] | Prospective | 0 - 14y | Sepsis or shock | 60 | 0.58 | | | |
| Joffre 2016[87] | Retrospective | 0 - 16y | Down's | 66.67 | 0.61 | | | |
| Muttath 2019[88] | Prospective | 0 - 16y | Sepsis or shock | 47.08 | 0.61 | | | |
| Misirlioglu 2018[89] | Retrospective | mean 5.2y | Whole PICU | 53.13 | 0.63 | | | |
| Appiah 2018[90] | Retrospective | 0 - 13y | Arrest | 58.25 | 0.63 | | | |

| Study | Design | Age | Population | % Male | F/M OR | Reported | Estimate | Adjustment |
|---|---|---|---|---|---|---|---|---|
| Celiny 2007[91] | Retrospective | 1m - 15y | Sepsis or shock | 66.67 | 0.67 | | | |
| Tong 2016[92] | Retrospective | <12y | Paramyxovirus Infection | 47.96 | 0.67 | | | |
| Cortina 2019[93] | Retrospective | median 5.6y | RRT | 52.8 | 0.69 | | | |
| Choi 2017[94] | Retrospective | mean 9.5y | AKI | 44.72 | 0.7 | | | |
| Basnet 2014[95] | Prospective | 0 - 16y | Whole PICU | 56.56 | 0.72 | | | |
| El-Mekkawy 2020[96] | Prospective | 0 - 16y | Whole PICU | 55.13 | 0.73 | | | |
| Ghani 2012[97] | Prospective | median 4.7m | Viral respiratory tract infections | 49.14 | 0.77 | | | |
| Sik 2019[98] | Retrospective | 0 - 18y | RRT | 42.86 | 0.77 | | | |
| Sial 2019[99] | Observation | 1 - 17y | Oncology | 61.02 | 0.8 | | | |
| Andersson 2019[100] | Retrospective | 0 - 18y | MOF | 57.14 | 0.8 | | | |
| Wong 2019[101] | Retrospective | 0 - 157y | Sepsis or shock | 44.83 | 0.81 | | | |
| Kim 2020[102] | Prospective | median 13y | Oncology | 56.36 | 0.81 | | | |
| Dewi 2020[103] | Retrospective | 0 - 18y | Whole PICU | 60.67 | 0.81 | | | |
| Chong 2015[104] | Retrospective | <16y | TBI/Head injury | 63.64 | 0.85 | | | |
| Naghib 2010[105] | Retrospective | Children | Long stay | 57.14 | 0.86 | | | |
| Patki 2017[106] | Prospective | 0 - 16y | Whole PICU | 57.86 | 0.86 | | | |
| Siddiqui 2018[107] | Retrospective | 0 - 16y | Whole PICU | 58.68 | 0.86 | | | |
| Proulx 1994[108] | Retrospective | <18y | MOF | 58.82 | 0.87 | | | |
| Lopez 2006[109] | Prospective | 0 - 18y | Whole PICU | 57.49 | 0.87 | | | |

| Study | Design | Age | Population | % Male | F/M OR | Reported | Estimate | Adjustment |
|---|---|---|---|---|---|---|---|---|
| Kim 2020[110] | Retrospective | 0 - 18y | RBC distribution width reading | 51.56 | 0.87 | | | |
| Tijssen 2020[111] | Retrospective | <18y | Transported to PICU | 58.28 | 0.87 | | | |
| Saboktakin 2020[112] | Prospective | 0 - 13y | Pneumonia/Resp tract infection | 57.92 | 0.88 | | | |
| Miklaszewska 2019[113] | Retrospective | mean 9.3y | RRT | 58.7 | 0.89 | | | |
| Ghuman 2013[114] | Retrospective | 2 -7y | Sepsis or shock | 52.7 | 0.9 | | | |
| Kanwaljeet 2015[115] | Prospective | <18y | Whole PICU | 56.74 | 0.9 | OR m/f | 1.1 | Age, Race/Ethnicity, Insurance type, Sepsis |
| Rusmawatiningtyas 2016[116] | Prospective | 0 - 17y | Sepsis or shock | 52.21 | 0.9 | | | |
| Selewski 2011[117] | Retrospective | median 19m | RRT | 59.29 | 0.91 | | | |
| Modem 2014[118] | Retrospective | median 9.5y | RRT | 41.05 | 0.91 | | | |
| Patel 2017[119] | Retrospective | 0 - 18y | Intoxication | 44.39 | 0.93 | | | |
| Hon 2017[120] | Retrospective | <12y | Whole PICU | 55.45 | 0.93 | | | |
| Kazantzi 2017[121] | Retrospective | median 60d | Pertussis infection | 32.26 | 0.94 | | | |
| Li 2019[122] | Prospective | 0 - 168m | RBC distribution width reading | 61.14 | 0.97 | | | |
| Khan 2020[123] | Prospective | 0 - 18y | Whole PICU | 53.51 | | OR m/f | 1.3 | Age, Race/Ethnicity, Primary diagnosis |
| Memon 2020[124] | Cross-sectional | mean 7y | Oncology | 66.67 | | OR m/f | 1.27 | Age, Inotropes, Mechanical ventilation |

F/M OR: This is the female/male OR we calculated from the crude numbers provided, Reported: This is the measure provided by the authors of the study, Adjustment: Any variables used for adjustment for the reported measure, AKI: Acute kidney injury, ARDS: Acute respiratory distress

| Study | Design | Age | Population | % Male | F/M OR | Reported | Estimate | Adjustment |
|---|---|---|---|---|---|---|---|---|
| syndrome, Arrest: Cardiorespiratory arrest, BG: Blood glucose, ECMO: Extra-corporeal membrane oxygenation, MOF: Multi organ failure, MV: Mechanical ventilation, RRT: Renal replacement therapy, TBI: Traumatic brain injury, SES: Socio-economic status | | | | | | | | |
