## Appendix 4 (Additional plots for some of the reported sub-populations for "Is there a sex difference in mortality rates in Paediatric Intensive Care Units: A Systematic Review"

Female/male mortality for PICU patients admitted with sepsis

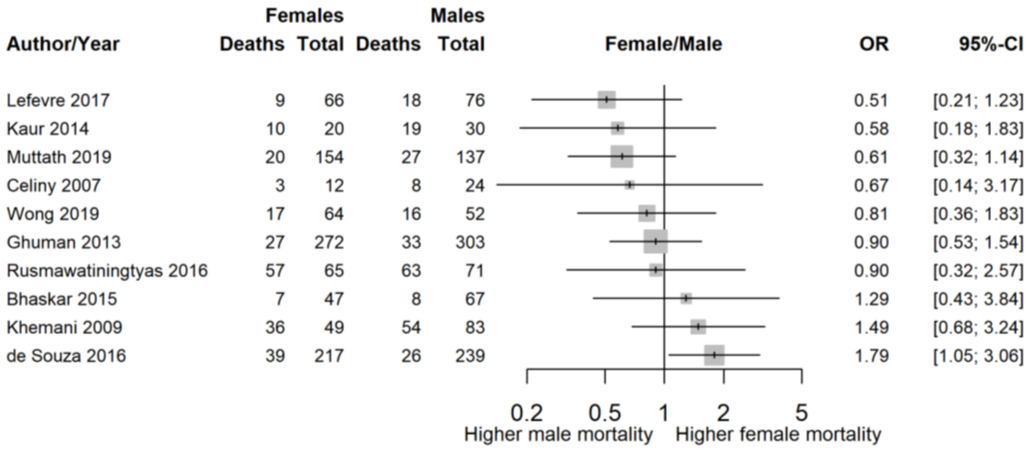


Female/male mortality for PICU patients admitted with renal replacement therapy

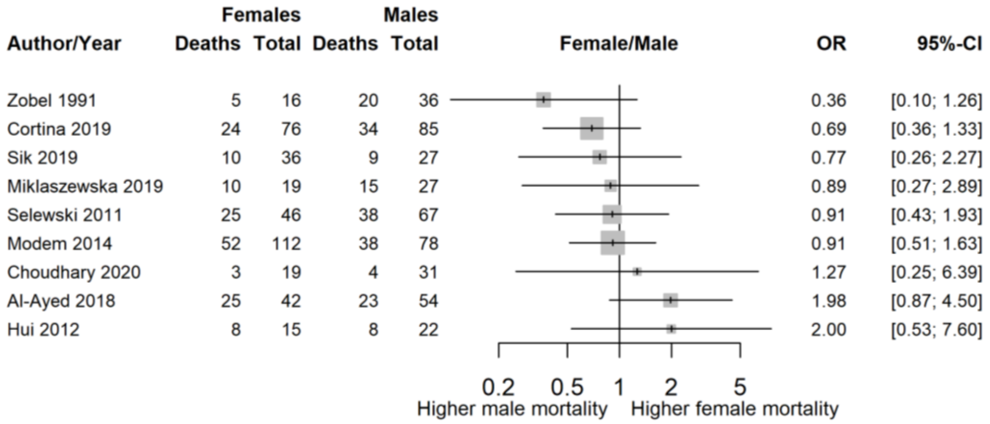

Female/male mortality for PICU patients admitted with bone marrow transplant

| | Females | | Males | | | |
|---|---|---|---|---|---|---|
| Author/Year | Deaths | Total | Deaths | Total | OR | 95%-CI |
| Lombel 2012 | 2 | 7 | 6 | 14 | 0.53 | [0.08; 3.76] |
| Raymakers-Janssen 2019 | 14 | 25 | 23 | 43 | 1.11 | [0.41; 2.98] |
| An 2016 | 3 | 9 | 5 | 20 | 1.50 | [0.27; 8.34] |
| Jacobe 2003 | 9 | 11 | 20 | 29 | 2.02 | [0.36; 11.34] |
| Rowan 2016 | 78 | 97 | 56 | 125 | 5.06 | [2.74; 9.34] |

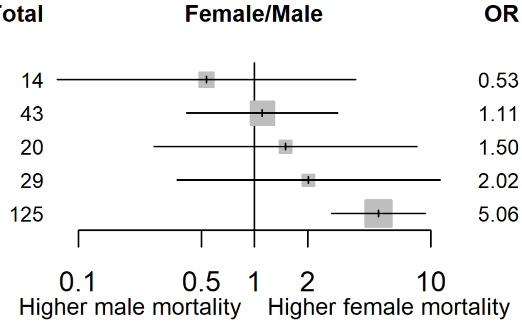


Female/male mortality for PICU patients admitted with oncological conditions

| | Females | | Males | | | |
|---|---|---|---|---|---|---|
| Author/Year | Deaths | Total | Deaths | Total | OR | 95%-CI |
| Dursun 2020 | 17 | 27 | 19 | 21 | 0.18 | [0.03; 0.93] |
| Sial 2019 | 9 | 23 | 16 | 36 | 0.80 | [0.28; 2.33] |
| Kim 2020 | 27 | 48 | 38 | 62 | 0.81 | [0.38; 1.75] |
| Sayed 2018 | 28 | 45 | 30 | 78 | 2.64 | [1.24; 5.61] |
| Ali 2016 | 136 | 220 | 82 | 330 | 4.90 | [3.38; 7.08] |

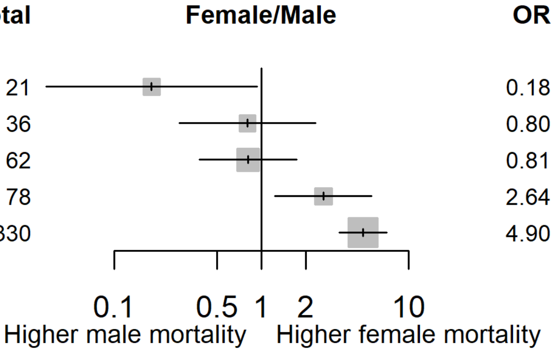

### Female/male mortality for PICU patients admitted with acute kidney injury

| Author/Year | Females Deaths | Females Total | Males Deaths | Males Total | OR | 95%-CI |
|---|---|---|---|---|---|---|
| Choi 2017 | 25 | 68 | 25 | 55 | 0.70 | [0.34; 1.44] |
| Alobaidi 2020 | 14 | 442 | 18 | 575 | 1.01 | [0.50; 2.06] |
| Krishnamurthy 2013 | 12 | 25 | 13 | 29 | 1.14 | [0.39; 3.32] |
| Ferreira 2020 | 44 | 188 | 52 | 246 | 1.14 | [0.72; 1.80] |
| Riyuzo 2017 | 11 | 30 | 15 | 47 | 1.24 | [0.47; 3.24] |

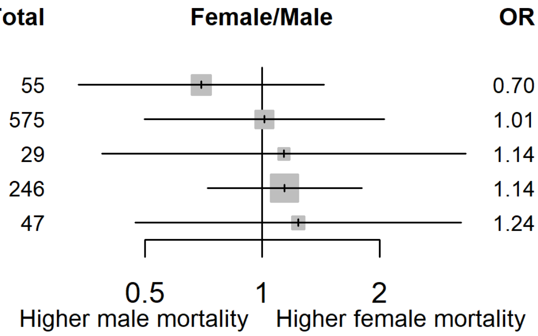


### Female/male mortality for PICU patients on mechanical ventilation

| Author/Year | Females Deaths | Females Total | Males Deaths | Males Total | OR | 95%-CI |
|---|---|---|---|---|---|---|
| Branco 2011 | 8 | 75 | 16 | 69 | 0.40 | [0.16; 0.99] |
| Sinitsky 2015 | 30 | 276 | 23 | 361 | 1.79 | [1.02; 3.16] |
| Shanmugham 2018 | 52 | 59 | 29 | 71 | 10.76 | [4.29; 27.00] |

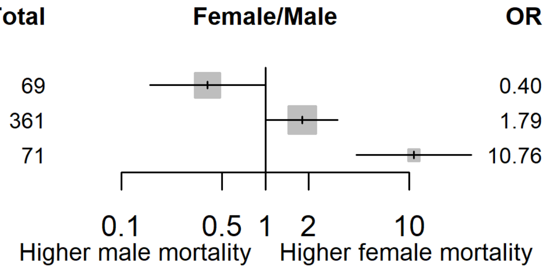
